# A Comprehensive Head-to-Head Comparison of Key Plasma Phosphorylated Tau 217 Biomarker Tests

**DOI:** 10.1101/2024.07.02.24309629

**Authors:** Noëlle Warmenhoven, Gemma Salvadó, Shorena Janelidze, Niklas Mattsson-Carlgren, Divya Bali, Anna Orduña Dolado, Hartmuth Kolb, Gallen Triana-Baltzer, Nicolas R. Barthélemy, Suzanne E. Schindler, Andrew J. Aschenbrenner, Cyrus A. Raji, Tammie L.S. Benzinger, John C. Morris, Laura Ibanez, Jigyasha Timsina, Carlos Cruchaga, Randall J. Bateman, Nicholas Ashton, Burak Arslan, Henrik Zetterberg, Kaj Blennow, Alexa Pichet Binette, Oskar Hansson

## Abstract

Plasma phosphorylated-tau 217 (p-tau217) is currently the most promising biomarkers for reliable detection of Alzheimer’s disease (AD) pathology. Various p-tau217 assays have been developed, but their relative performance is unclear. We compared key plasma p-tau217 tests using cross-sectional and longitudinal measures of amyloid-β (Aβ)-PET, tau-PET, and cognition as outcomes, and benchmarked them against cerebrospinal fluid (CSF) biomarker tests.

Samples from 998 individuals (mean[range] age 68.5[20.0-92.5], 53% female) from the Swedish BioFINDER-2 cohort were analyzed. Plasma p-tau217 was measured with mass spectrometry (MS) assays (the ratio between phosphorylated and non-phosphorylated [%p-tau217_WashU_]and p-tau217_WashU_) as well as with immunoassays (p-tau217_Lilly_, p-tau217_Janssen_, p-tau217_ALZpath_). CSF biomarkers included p-tau217_Lilly_, and the FDA-approved p-tau181/Aβ42_Elecsys_ and p-tau181_Elecsys_.

All plasma p-tau217 tests exhibited high ability to detect abnormal Aβ-PET (AUC range: 0.91-0.96) and tau-PET (AUC range: 0.94-0.97). Plasma %p-tau217_WashU_ had the highest performance, with significantly higher AUCs than all the immunoassays (*P*_diff_<0.007). For detecting Aβ-PET status, %p-tau217_WashU_ had an accuracy of 0.93 (immunoassays: 0.83-0.88), sensitivity of 91% (immunoassays: 84-87%), and a specificity of 94% (immunoassays: 85-89%). Among immunoassays, p-tau217_Lilly_ and plasma p-tau217_ALZpath_ had higher AUCs than plasma p-tau217_Janssen_ for Aβ-PET status (*P*_diff_<0.006), and p-tau217_Lilly_ outperformed plasma p-tau217_ALZpath_ for tau-PET status (*P*_diff_=0.025). Plasma %p-tau217_WashU_ exhibited higher associations with all PET load outcomes compared to immunoassays; baseline Aβ-PET load (R^2^: 0.72; immunoassays: 0.47-0.58; P_diff_<0.001), baseline tau-PET load (R^2^: 0.51; immunoassays: 0.38-0.45; P_diff_<0.001), longitudinal Aβ-PET load (R^2^: 0.53; immunoassays: 0.31-0.38; P_diff_<0.001) and longitudinal tau-PET load (R^2^: 0.50; immunoassays: 0.35-0.43; P_diff_<0.014). Among immunoassays, plasma p-tau217_Lilly_ was more strongly associated with Aβ-PET load than plasma p-tau217_Janssen_ (*P*_diff_<0.020) and with tau-PET load than both plasma p-tau217_Janssen_ and plasma p-tau217_ALZpath_ (all *P*_diff_<0.010). Plasma %p-tau217 also correlated more strongly with baseline cognition (Mini-Mental State Examination[MMSE]) than all immunoassays (R^2^ %p-tau217_WashU_: 0.33; immunoassays: 0.27-0.30; *P*_diff_<0.024). The main results were replicated in an external cohort from Washington University in St Louis (*n* =219). Finally, p-tau217_Nulisa_ showed similar performance to other immunoassays in subsets of both cohorts.

In summary, both MS-and immunoassay-based p-tau217 tests generally perform well in identifying Aβ-PET, tau-PET, and cognitive abnormalities, but %p-tau217_WashU_ performed significantly better than all the examined immunoassays. Plasma %p-tau217 may be considered as a stand-alone confirmatory test for AD pathology, while some immunoassays might be better suited as triage tests where positive results are confirmed with a second test.

## Introduction

Detection of Alzheimer’s disease (AD) pathology, i.e., amyloid-β (Aβ) plaques and hyperphosphorylated tau aggregates, with accurate blood tests is of high interest both for research and clinical trials, and clinical implementation is already underway.^1–3^ For optimal clinical management of AD, it is crucial to ensure a timely and accurate diagnosis in a scalable and cost-effective manner. This is becoming even more important with the launch of disease-modifying therapies in several countries.^1,4,5^ Tau phosphorylated at threonine 217 (p-tau217) has emerged as a leading plasma biomarker of AD pathology along the disease continuum ranging from pre-symptomatic to symptomatic disease stages.^1,3,6–10^ Several p-tau217 assays have been developed using a variety of methods. The performance of these different assays may differ depending on analytical factors (e.g., the specific antibodies used), or the measurement techniques (e.g., mass spectrometry [MS] versus immunoassays). A comparison of the different assays is needed to determine to what extent different assays provide an alternative to established cerebrospinal fluid (CSF) and imaging markers. Previously, we showed that MS-based measures of p-tau217 performed the best out of a variety of p-tau217 and p-tau181 plasma assays to predict CSF Aβ-status in 135 individuals with mild cognitive impairment (MCI).^8^ It has also been shown that several p-tau217 immunoassays perform similarly when predicting Aβ-status.^11–14^ Nonetheless, it is not yet clear whether different p-tau217 assays perform similarly in relation to other key measures of AD, such as cross-sectional and longitudinal measures of Aβ-PET load, tau-PET load, and cognition, and how they compare to well-established CSF markers in these contexts. Further, it is important to determine which plasma p-tau217 tests fulfill the proposed minimal requirements for use in clinical practice either as i) a stand-alone confirmatory test, or ii) as a triage test where positive or intermediate results are confirmed with a higher performing test such as CSF or PET.^15^

To this end, we conducted a head-to-head study comparing the performance and accuracy of four different plasma p-tau217 assays, using both high-performing MS- and immunoassay-based tests in a large Swedish sample covering the full spectrum of AD (*n* = 998) with sensitivity analyses performed in cognitively unimpaired (CU) and cognitively impaired (CI) individuals separately. The blood tests were compared in their abilities to discriminate normal versus abnormal Aβ- and tau-PET status, and for continuous associations with baseline levels and the rate of change in Aβ- and tau-PET values, and cognitive test scores. Plasma biomarker performances were also compared to relevant CSF tests, i.e., the FDA-approved p-tau181/Aβ42 ratio and p-tau181 tests, and p-tau217. Finally, the results were replicated using plasma biomarkers in an external American cohort (*n* = 219). We hypothesized that the best plasma p-tau217 tests would be on par with the CSF tests, supporting a possible substitution of CSF tests with plasma biomarkers for AD.

## Materials and methods

### Participants

#### BioFINDER-2 cohort

The study population consisted of participants from the prospective Swedish BioFINDER-2 study (NCT03174938), recruited between 2017-2022, from the Skåne University Hospital and Ängelholm Hospital. Participants were either cognitively normal (*n* = 375), had subjective cognitive decline (SCD) (*n* = 139), mild cognitive impairment (MCI) (*n* = 256), or dementia due to AD (*n* = 131), or due to other causes (frontotemporal dementia, dementia with Lewy bodies, Parkinson’s disease dementia, vascular dementia or unspecified dementia) (*n* = 97). Overall exclusion criteria included: i) presence of systemic illness preventing study participation; ii) significant neurological or psychiatric diseases; iii) current alcohol or substance abuse; iv) unwilling to undergo imaging or lumbar puncture. All participants were required to be proficient in Swedish. Cognitively normal and SCD participants formed the group of CU individuals (*n* = 514). The inclusion criteria here included not meeting MCI or dementia criteria (for further details see Salvadó et al., 2024). Participants with MCI exhibited significant cognitive decline on at least one domain from a neuropsychological test battery, defined as performing below 1.5 standard deviation below the normative score.^16^ AD dementia was diagnosed based on DSM-5 criteria and further confirmed with Aβ biomarkers based on the NIA-AA criteria for AD.^17^ Patients with MCI or dementia formed the group of CI individuals (*n* = 484). For this study, participants were only included if they had all plasma and CSF biomarkers available at baseline (*n* = 998).^18,19^ The BioFINDER-2 study was approved by the Swedish Ethical Review Authority. Each participant and/or their relatives provided informed consent. The study adhered to guidelines outlines in the WMA Declaration of Helsinki and the Department of Health and Human Services Belmont Report.

#### Knight ADRC cohort

The Knight ADRC cohort is composed of community-dwelling volunteers enrolled in aging studies in Washington University in St. Louis. All participants underwent comprehensive clinical assessments including neurological examinations and cognitive testing and were included if they had all fluid biomarkers available (*n* = 219).^20^ Participants with a Clinical Dementia Rating (CDR)=0 were classified as cognitively unimpaired (*n* = 195) and those with a CDR>0 were classified as cognitively impaired (*n* = 27).^21^ All participants gave written informed consent and ethical approval was granted by the Washington University Human Research Protection Office (protocol: 201109100).

### Plasma biomarkers

Blood was collected in K2-EDTA-plasma tubes and centrifuged at 2000*g*, +4°C for 10 minutes. Plasma was aliquoted into 1.5-ml polypropylene tubes (1 ml per tube) and stored at -80°C. Plasma %p-tau217_WashU_ and plasma p-tau217_WashU_ were derived from non-phosphorylated tau217 and p-tau217 concentrations were measured with a liquid chromatography tandem high-resolution mass spectrometry (LC-MS/HRMS) method developed at Washington University (WashU).^7,22^ The plasma %p-tau217_WashU_ was determined by dividing the level of tau phosphorylated at residue 217 by the concentration of the non-phosphorylated version of the same tau peptide. Plasma p-tau217_Lilly_ concentrations were measured with the Meso Scale Discovery immunoassay developed by Lilly Research Laboratories.^23–25^ Plasma p-tau217_Janssen_ concentrations were measured using a Single molecule arrays (Simoa) immunoassay developed by Johnson & Johnson Innovative Medicine, formerly Janssen R&D^12,26,27^, which is commercially available at Quanterix as LucentAD p217 assay. Plasma p-tau217_ALZpath_ was measured with the ALZpath pTau217 assay and is commercially available.^28^ More detailed descriptions regarding the assays can be found in Supplementary materials. In the Knight ADRC cohort, plasma biomarkers were measured with the same assays as in BioFINDER-2, except that the Janssen and ALZpath assays were not available. For a subset of participants, measurements with the multiplex NULISA pTau-217 Assay (p-tau217_NULISA_)^29^ were available in BioFINDER-2 (*n* = 463) and the Knight ADRC cohort (*n* = 97).

### CSF biomarkers

CSF was obtained through lumbar puncture during the same session as blood collection and stored at -80°C polypropylene tubes. Samples were handled in agreement with the suggested protocol.^30^ CSF p-tau217_Lilly_ concentrations were measured with the Meso Scale Discovery immunoassay developed by Lilly Research Laboratories. CSF p-tau181, Aβ40 and Aβ42 concentrations were measured with the Roche Elecsys CSF immunoassays on a fully automated Cobas instruments (Roche Diagnostics International Ltd., Rotkreuz, Switzerland).^31^ The ratio between CSF p-tau181 and Aβ42 (CSFp-tau181/Aβ42) is approved by the FDA to detect Alzheimer’s pathology in cognitively impaired individuals. Participants were categorized as being CSF Aβ-positive based on a CSF Aβ42/Aβ40 threshold below 0.075, determined with Gaussian mixture modelling (GMM).^24,32^ In the Knight ADRC cohort, CSF was collected at the same time as blood draw. CSF Aβ42, Aβ40 and p-tau181 were measured with the Lumipulse G1200 automated immunoassay platform (Fujirebio). The Aβ42/40 ratio from Lumipulse is also FDA-approved.

### PET

PET scans were performed at Skåne University Hospital in Lund, Sweden. Aβ-PET images were acquired 90-110 min after the injection of ∼185 MBq [^18^F]flutemetamol in participants without dementia (*n* =698). The standardized uptake value ratios (SUVR) were created with the whole cerebellum as reference region. Aβ-PET positivity was defined as having a SUVR in the neocortical meta region of interest (ROI) of ≥1.033, defined through GMM.^33^ A subset of participants (*n* = 448) underwent Aβ-PET longitudinally for an average follow-up of 3.63 ± 2.30 years. Almost all participants (*n* = 961) had tau-PET data available. Images were acquired 70-90 min post injection of ∼370 MBq of [^18^F]RO948. Tau-PET positivity was characterized as ≥1.362 SUVR in the temporal-meta ROI with the inferior cerebellar cortex as reference region.^34,35^ A subset of participants (*n =* 524) had follow-up tau-PET data available over the course of 3.46 ± 2.09 years. Further details have been described elsewhere.^24,36^ The Swedish Medical Products Agency and the local Radiation Safety Committee at Skåne University Hospital, Sweden, approved the PET imaging. In the Knight ADRC cohort, PET-scans were obtained within one year of plasma and CSF collection. Aβ-PET was performed with either tracer [^18^F]florbetapir (AV45) or [^11^C]Pittsburgh Compound B (PiB).^20^ The mean cortical SUVR was calculated based on the averaged signal of neocortical ROIs using cerebellar gray matter as the reference regions. Centiloids were derived from SUVR values and a cutoff of ³37 Centiloids was defined for Aβ-PET positivity.^37^

### Cognition

We focused on two cognitive measures. First, we included the MMSE^38^, which measured global cognitive function, with a range of zero to thirty, in which zero is the worst possible score. Second, we included a modified version of the Preclinical Alzheimer Cognitive Composite (mPACC), which measures different aspects of cognition associated with early stages of cognitive decline, in which lower scores indicate more cognitive impairment. The mPACC included z-scores on animal fluency, MMSE, Symbol Digit Modalities Test and Alzheimer’s Disease Assessment Scale delayed recall scores (counted twice).^39^ Dementia patients were excluded from mPACC analyses (*n* = 228). In the Knight ADRC cohort, we used a global cognitive composite as previously described.^40^

### Statistical analyses

Statistical analyses were carried out in R (version 4.3.2). Fluid biomarkers were log_10_-transformed (except p-tau217_NULISA_) and z-scored using CU Aβ− individuals as reference group. Correlations between biomarkers were assessed with scaled linear regression models. Abilities to discriminate between Aβ- or tau-PET positive or negative participants were assessed with receiver operating characteristic (ROC) curve analysis (‘pROC’ package). The area under the curve (AUC) of two ROC curves was compared with bootstrapping (1000 iterations). In brief: significant differences in test metrics (e.g. AUC) were assessed by analyzing the generated distribution of the bootstrapped differences. Sensitivity, specificity, positive predictive values (PPV) and negative predictive values (NPV) were calculated using the threshold derived from the optimization of the Youden’s index (‘pROC’ package). In a complementary analysis, for each biomarker, we grouped participants into PET-negative and further split the PET-positive group into quartiles. Wilcoxon signed-rank tests were used to compare differences between neighboring quartiles. We also looked at the associations with continuous measures of PET SUVR and cognitive performance at baseline as well as their rates of change. For the linear regression models, we included Aβ-PET, tau-PET, MMSE score and mPACC as outcomes with the plasma or CSF biomarkers as predictors (1 model per biomarker and outcome). For each outcome, the adjusted R^2^ of each biomarker model was compared using the same bootstrapping method (same sample size, 1000 iterations). Linear mixed models were used to obtain the individual rate-of-change (‘lme4 package’) from longitudinal Aβ-PET, tau-PET, MMSE score and mPACC, using random intercepts and random time slopes. These slopes were then used as the outcome in a linear regression model with the fluid biomarkers as predictors. Again, we compared the adjusted R^2^ of each biomarker model with bootstrapping (1000 iterations). The same analyses were performed in the Knight ADRC cohort, with the available outcomes: Aβ-PET status or SUVR and a global cognitive composite. The AUC models and linear regression models were all adjusted for age and sex. Cognition models were additionally adjusted for years of education. The p-values were adjusted for multiple comparisons using the Benjamini Hochberg false discovery rate (FDR) method, applied across all comparisons per model, for each outcome. Hence, all reported *p*-values are FDR-corrected. Two-sided *p* <0.05 was considered as statistically significant. All main analyses were performed including all participants. Analyses were then repeated only in CU and only in CI as sensitivity analyses.

### Data availability

Anonymized data will be shared by request from a qualified academic investigator for the sole purpose of replicating procedures and results presented in the article and if data transfer agrees with EU legislation on the general data protection regulation and decisions by the Ethical Review Board of Sweden and Region Skåne, which should be regulated in a material transfer agreement.

## Results

### Demographics and correlations between biomarkers

Plasma and CSF concentrations were assessed in 998 individuals in the BioFINDER-2 study, with a mean age of 68.5 (12.1) and 53% women. In total, 52% (*n* = 514) was considered as CU, and 48% (*n* = 484) as CI (see **Table 1** for demographic information). When correlating the different plasma tests with each other, the two MS plasma methods, as expected, had the highest correlation with an R^2^ of 0.92, whereas other correlations ranged between 0.61 and 0.80 (**Fig. 1, Supplementary Fig. 1**). It must be noted that as p-tau217 is the numerator for %p-tau217, these measures are not independent.

**Figure 1.**
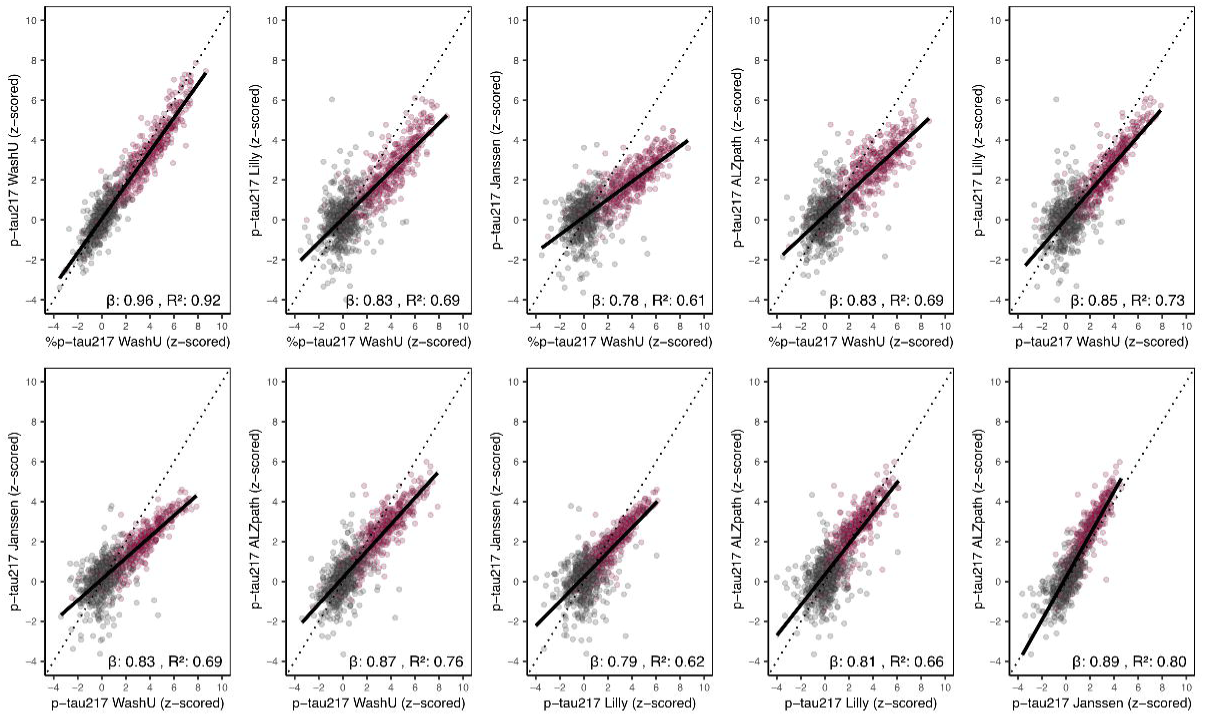
Correlations between plasma p-tau217 biomarkers. The reported betas are standardized and across the full sample. Z-scores were based on cognitively unimpaired, CSF Aβ− participants (*n* = 364) as the reference group. *Abbreviations*: Aβ, amyloid-beta.

### Classification of A**β**-PET status

First, we assessed how well the different assays could identify individuals with an abnormal Aβ-PET status using ROC analyses (**Fig. 2A**). Plasma %p-tau217_WashU_ performed significantly better than the other plasma biomarkers with an AUC of 0.96 (all *P*_diff_ < 0.001). Among the immunoassays, plasma p-tau217_Lilly_ (AUC: 0.94) and plasma p-tau217_ALZpath_ (AUC: 0.93) both had significantly higher AUCs than plasma p-tau217_Janssen_ (AUC: 0.91, all *P*_diff_ < 0.016) (**Supplementary Table 1**). When comparing to CSF tests, no significant difference was observed between plasma %-ptau217_WashU_ (AUC: 0.96) and CSF p-tau217_Lilly_ (AUC: 0.95, *P*_diff_ = 0.074, **Fig. 2A**). Plasma %p-tau217_WashU_ (AUC: 0.96) performed significantly better than CSF p-tau181/Aβ42_Elecsys_ (AUC: 0.94, *P*_diff_ = 0.007), whereas plasma p-tau217_WashU_, plasma p-tau217_Lilly_ and plasma p-tau217_ALZpath_ performed similarly to CSF p-tau181/Aβ42_Elecsys_ (all *P*_diff_ > 0.377). Plasma p-tau217_Janssen_ (AUC: 0.91) was inferior to CSF p-tau181/Aβ42_Elecsys_ (AUC: 0.94, *P*_diff_ = 0.012). CSF p-tau181_Elecsys_ had the lowest AUCs of all biomarker tests (either plasma or CSF) (**Fig. 2A** and **Supplementary Table 1)**.

**Figure 2.**
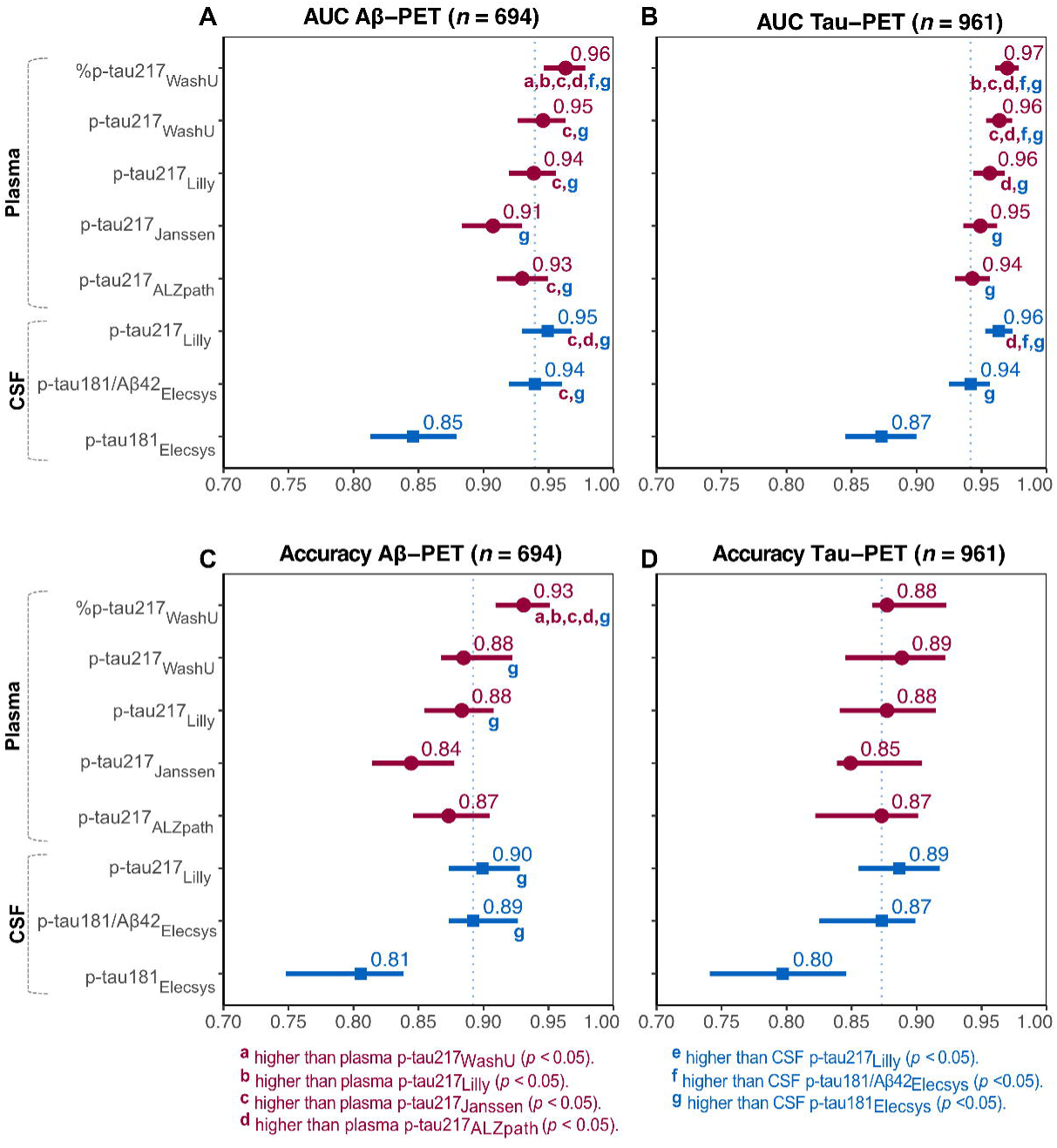
Comparisons of AUC and accuracy between plasma p-tau217 biomarkers for Aβ-PET and tau-PET status. Squares represent the AUC or accuracy, and bars represent 95%CI. The dashed line is drawn at CSF p-tau181/Aβ42_Elecsys_ to facilitate comparing the other tests to the current approved FDA-approved test. Significant differences between assays were assessed through bootstrapping and all p-values were FDR-corrected. Models were corrected for age and sex. *Abbreviations*: Aβ, amyloid-beta; CI, confidence interval; FDR, false discovery rate.

Next, we studied the agreement between fluid biomarker tests and Aβ-PET status (**Fig. 2C** and **Table 2**). Plasma %p-tau217_WashU_ exhibited significantly higher accuracy compared to all other plasma biomarker tests (%p-tau217, 0.93; immunoassays, 0.83-0.88 , all *P*_diff_ < 0.025) (**Fig. 2C**). No significant differences between the accuracies of the plasma immunoassays were observed (all *P*_diff_ > 0.183). Further, plasma %p-tau217_WashU_ had a sensitivity of 91% (immunoassays: 84-87%) and a specificity of 94% (immunoassays: 85-89%) for detecting elevated Aβ-PET status (**Table 2**).

### Classification of tau-PET status

Next, we examined how well the different diagnostic tests identified an abnormal tau-PET status using ROC analyses (**Fig. 2B and Supplementary Table 1**). Plasma %p-tau217_WashU_ (AUC: 0.97) performed significantly better than all the plasma p-tau217 immunoassays (AUC: 0.94-0.96, all *P*_diff_ < 0.007). Among the immunoassays, plasma p-tau217_Lilly_ (AUC: 0.96) and plasma p-tau217_Janssen_ (AUC: 0.95) did not perform differently from each other (*P*_diff_ = 0.162;). However, plasma p-tau217_Lilly_ (AUC: 0.96) had a significantly higher AUC than plasma p-tau217_ALZpath_ (AUC: 0.94, *P*_diff_ = 0.025). When comparing to CSF tests, plasma %p-tau217_WashU_, plasma p-tau217_WashU_ and CSF p-tau217_Lilly_ had significantly higher AUCs (0.96-0.97) than CSF p-tau181/Aβ42_Elecsys_ (AUC: 0.94, all *P*_diff_ < 0.009), but plasma p-tau217_Janssen_ and ptau217_ALZpath_ (AUC: 0.94-0.95) performed comparable to CSF p-tau181/Aβ42_Elecsys_ (all *P*_diff_ > 0.404). CSF p-tau181_Elecsys_ had the lowest AUC (0.87), and all other assays (either from plasma or CSF) were superior.

When the accuracies were compared, no significant differences were observed between any of the fluid biomarkers, which ranged between 0.80 and 0.89 (**Fig. 2D**). More information regarding sensitivity and specificity can be found in **Table 2**.

### Distinguishing between PET quantiles

To better understand how the different fluid biomarkers tracked changes in the levels of the AD proteinopathies, we examined the levels of each fluid biomarker test between different PET stages (PET-negative participants in the “Negative” group and PET-positive participants further split into quartiles [Q1-Q4]). When grouping individuals by Aβ-PET SUVR, the CSF measures plateaued in individuals with the highest levels of Aβ pathology (between Q3 and Q4), but the plasma biomarkers continued to increase with higher PET load (**Fig. 3A**). For tau-PET, more differences were observed. Plasma %p-tau217_WashU_, plasma p-tau217_WashU_ and plasma p-tau217_Lilly_ did not plateau at later stages of tau-PET positivity and continued to increase with higher PET loads. Plasma p-tau217_Janssen_ did not show a significant difference when comparing Q2 and Q3 stage but did for the other stages. Plasma p-tau217_ALZpath_ only showed a significant difference when comparing the tau-PET negative group to Q1, and Q2 and Q2, but was not different for the other quantiles. The CSF Elecsys measures (p-tau181/Aβ42 and p-tau181) plateaued after distinguishing between the negative group and Q1, but CSF p-tau217_Lilly_ did not (**Fig. 3B**).

**Figure 3.**
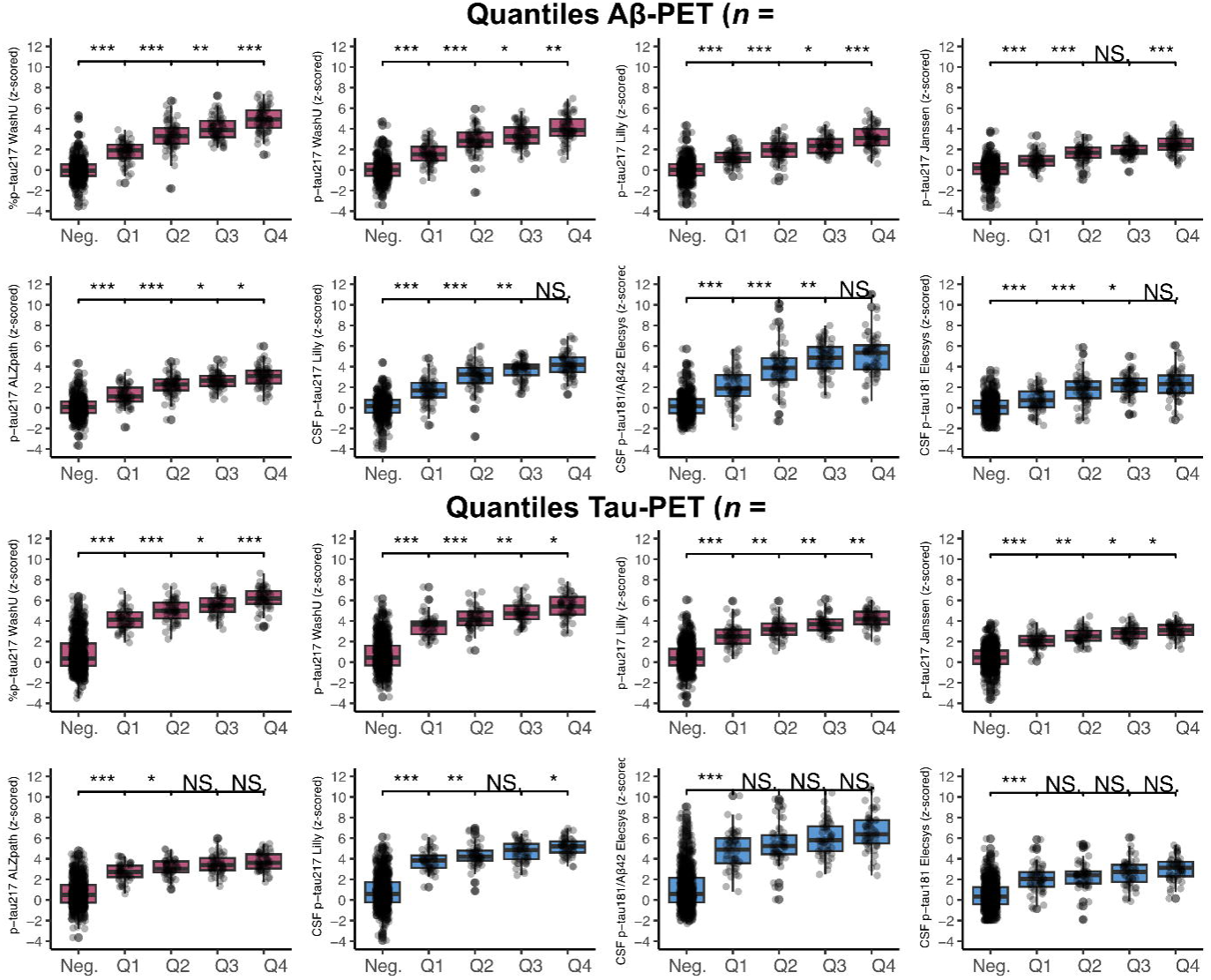
Quantile grouping for Aβ-PET and tau-PET. Boxes show the interquartile range and the horizontal lines represent the medians. Negative participants were defined as falling below the pre-defined cut-offs (Aβ-PET: <1.033; tau-PET: <1.362). Neighboring quantiles were compared with Wilcoxon signed-rank tests. *: p-value < 0.05. **: p-value < 0.01. ***: p-value < 0.001. *Abbreviations:* Aβ, amyloid-beta.

### Associations with A**β**-PET load

Next, we examined the associations between fluid biomarker test outcomes at baseline and i) Aβ-PET load at baseline, and ii) the rate of change of Aβ-PET load during follow-up. The results are given in **Fig. 4A** (cross-sectional analyses) and **Fig. 4C** (longitudinal analyses) with additional information in **Supplementary Tables 2 & 3** and **Supplementary Fig. 2 & 3.** Results across the different assays were very consistent between the two types of analyses. Overall, plasma %p-tau217_WashU_ showed the strongest association with Aβ-PET values, performing significantly better than all the plasma p-tau217 immunoassays with an R^2^ of 0.72 in the cross-sectional analysis (immunoassays: 0.47-0.58; all *P*_diff_ < 0.001), and with an R^2^ of 0.53 in the longitudinal analysis (immunoassays: 0.31-0.38; all *P*_diff_ < 0.005) . Among the immunoassays, plasma p-tau217Lilly (R^2^cross-sectional: 0.58, R^2^longitudinal: 0.38) and p-tau217ALZpath (R^2^cross-sectional: 0.56, R^2^_longitudinal_: 0.37) had a significantly higher R^2^ than plasma p-tau217_Janssen_ (R^2^_cross-sectional_: 0.47, R^2^_longitudinal_: 0.31) in both the cross-sectional and longitudinal analyses (all *P*_diff_ < 0.020) and did not differ from each other. When comparing plasma p-tau217 tests to CSF tests, plasma %p-tau217_WashU_ (R^2^_cross-sectional_: 0.72, R^2^_longitudinal_: 0.53) performed significantly better than CSF p-tau217_Lilly_ (R^2^_cross-sectional_: 0.65, R^2^ : 0.46) in both the cross-sectional and longitudinal analyses (all *P*_diff_ < 0.034) and than CSF p-tau181/Aβ42_Elecsys_ (R^2^_cross-sectional_: 0.62, R^2^ : 0.48) at the cross-sectional level (*P*_diff_ = 0.001). CSF p-tau217_Lilly_, however, performed significantly better than plasma p-tau217_Lilly_, plasma p-tau217_Janssen_ and p-tau217_ALZpath_ in both the cross-sectional and longitudinal analyses (all *P*_diff_ <0.021). CSF p-tau181/Aβ42_Elecsys_ performed better than p-tau217_Janssen_ in cross-sectional and longitudinal analyses (*P*_diff_ < 0.012) and p-tau217_ALZpath_ (R^2^: 0.37) in the longitudinal analyses (*P*_diff_ = 0.005). In the cross-sectional analyses, all plasma and CSF assays had stronger associations with Aβ-PET values than CSF p-tau181_Elecsys_ (R^2^ : 0.38, R^2^ : 0.25). This was replicated in the longitudinal analyses, except for plasma p-tau217_Janssen_.

**Figure 4.**
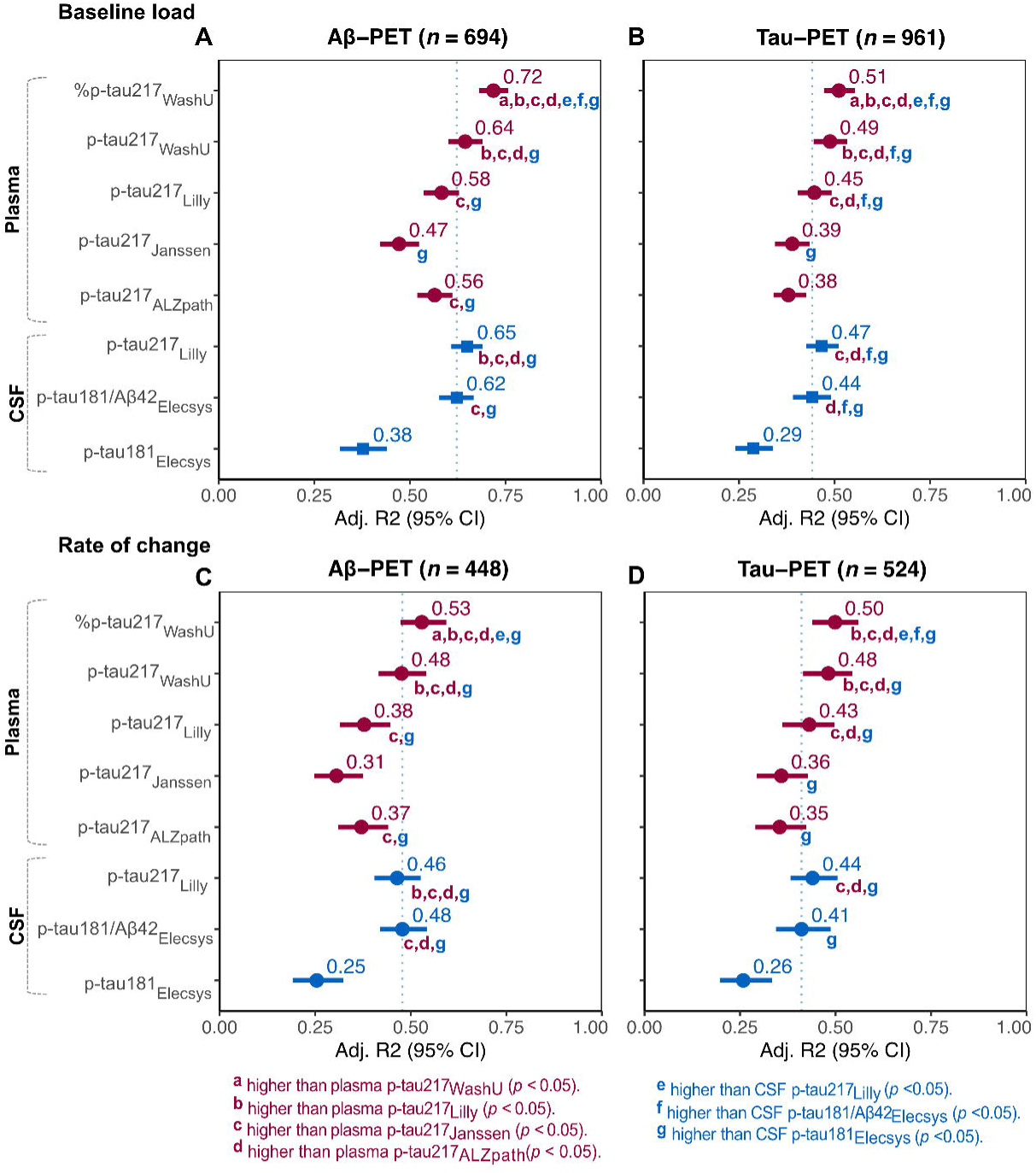
R^2^ comparisons with Aβ-PET and tau-PET load as outcome. Squares represent the AUC or accuracy, and bars represent 95%CI. The dashed line is drawn at CSF p-tau181/Aβ42_Elecsys_, to facilitate comparing the other tests to the current approved FDA-approved test. Significant differences between assays were assessed through bootstrapping and all p-values were FDR-corrected. Cross-sectional: PET SUVR ∼ biomarker + age + sex. Longitudinal: individual rate of change in PET SUVR ∼ biomarker + age + sex. *Abbreviations*: Aβ, amyloid-beta; CI, confidence interval; FDR, false discovery rate.

### Associations with tau-PET load

We then examined the associations between biomarker performance and tau-PET load. The results are given in **Fig. 4B** (cross-sectional analyses) and **Fig. 4D** (longitudinal analyses) with additional information in **Supplementary Tables 2 & 3** and **Supplementary Fig. 4 & 5.** Results were similar across both the cross-sectional and longitudinal analyses. Plasma %p-tau217_WashU_ performed significantly better than all the plasma p-tau217 immunoassays with an R^2^ of 0.51 in the cross-sectional analysis (immunoassays: 0.38-0.45; all *P*_diff_ < 0.001), and with an R^2^ of 0.50 in the longitudinal analysis (immunoassays: 0.35-0.43; all *P*_diff_ < 0.014). Among the plasma immunoassays, plasma p-tau217_Lilly_ (R^2^_cross-sectional_: 0.45, R^2^_longitudinal_: 0.43) performed significantly better than p-tau217_Janssen_(R^2^_cross-sectional_: 0.39, R^2^_longitudinal_: 0.36) and p-tau217_ALZpath_(R^2^_cross-sectional_: 0.38, R^2^_longitudinal_: 0.35) in both cross-sectional (all *P*_diff_ < 0.001) and longitudinal analyses (all *P*_diff_ < 0.019). Plasma p-tau217_Janssen_ and plasma p-tau217_ALZpath_ did not differ from each other in either analysis.

When comparing plasma p-tau217 tests to CSF tests, we found that plasma %p-tau217_WashU_ (R^2^_cross-sectional_: 0.51, R^2^_longitudinal_: 0.50) had a significantly higher R^2^ than both CSF p-tau217_Lilly_ (R^2^cross-sectional: 0.47, R^2^longitudinal: 0.44) and CSF p-tau181/Aβ42Elecsys (R^2^cross-sectional: 0.44, R^2^_longitudinal_: 0.41) in both the cross-sectional (all *P*_diff_ < 0.015) and longitudinal analyses (all *P*_diff_ < 0.043). However, CSF p-tau217_Lilly_ (R^2^_cross-sectional_: 0.47, R^2^_longitudinal_: 0.44) performed better than both p-tau217_Janssen_ (R^2^_cross-sectional_: 0.39, R^2^_longitudinal_: 0.36) and plasma p-tau217_ALZpath_ in both the cross-sectional (all *P*_diff_ < 0.001) and longitudinal analyses (R^2^_cross-sectional_: 0.38, R^2^_longitudinal_: 0.35, all *P*_diff_ < 0.017). Further, CSF p-tau181/Aβ42_Elecsys_ performed better than plasma p-tau217_ALZpath_ in the cross-sectional analysis (*P*_diff_ = 0.037). CSF p-tau181_Elecsys_ performed worse than all other tests.

### Associations with cognition

We next examined the biomarker associations with cognition cross-sectionally. The results were generally similar for both cross-sectional analyses of MMSE (**Fig. 5A**) and mPACC (**Fig. 5B**) and further information are given in **Supplementary Table 2** and **Supplementary Fig. 6 & 7**. Plasma %p-tau217_WashU_ had the largest cross-sectional associations with cognitive measures with an R^2^ of 0.33 for MMSE and 0.37 for the mPACC when controlling for age, sex and years of education. Plasma %p-tau217_WashU_ showed significantly larger associations with MMSE compared to all plasma p-tau217 immunoassays (R^2^: 0.27-0.30, all *P*_diff_ < 0.024), and larger associations with mPACC than plasma p-tau217_Lilly_ (R^2^: 0.33) and plasma p-tau217_Janssen_ (R^2^:0.33, all *P*_diff_ < 0.024). When comparing plasma and CSF tests, plasma %-tau217_WashU_ was equal or better compared to the CSF tests. CSF p-tau217_Lilly_ (R^2^: 0.34) exhibited stronger associations with MMSE than all plasma immunoassays (all *P*_diff_ < 0.042). Further, CSF p-tau181/Aβ42_Elecsys_ (R^2^: 0.32) exhibited stronger associations with MMSE than plasma p-tau217_Janssen_ (R^2^: 0.27) and p-tau217_ALZpath_ (R^2^: 0.27, all *P*_diff_ < 0.030). With the mPACC as the outcome, both CSF p-tau217_Lilly_ (R^2^: 0.37) and CSF p-tau181/Aβ42_Elecsys_ (R^2^: 0.38) exhibited stronger associations than plasma p-tau217_Lilly_ (R^2^: 0.33) and p-tau217_Janssen_ (R^2^: 0.33, all *P*_diff_ < 0.043).

**Figure 5.**
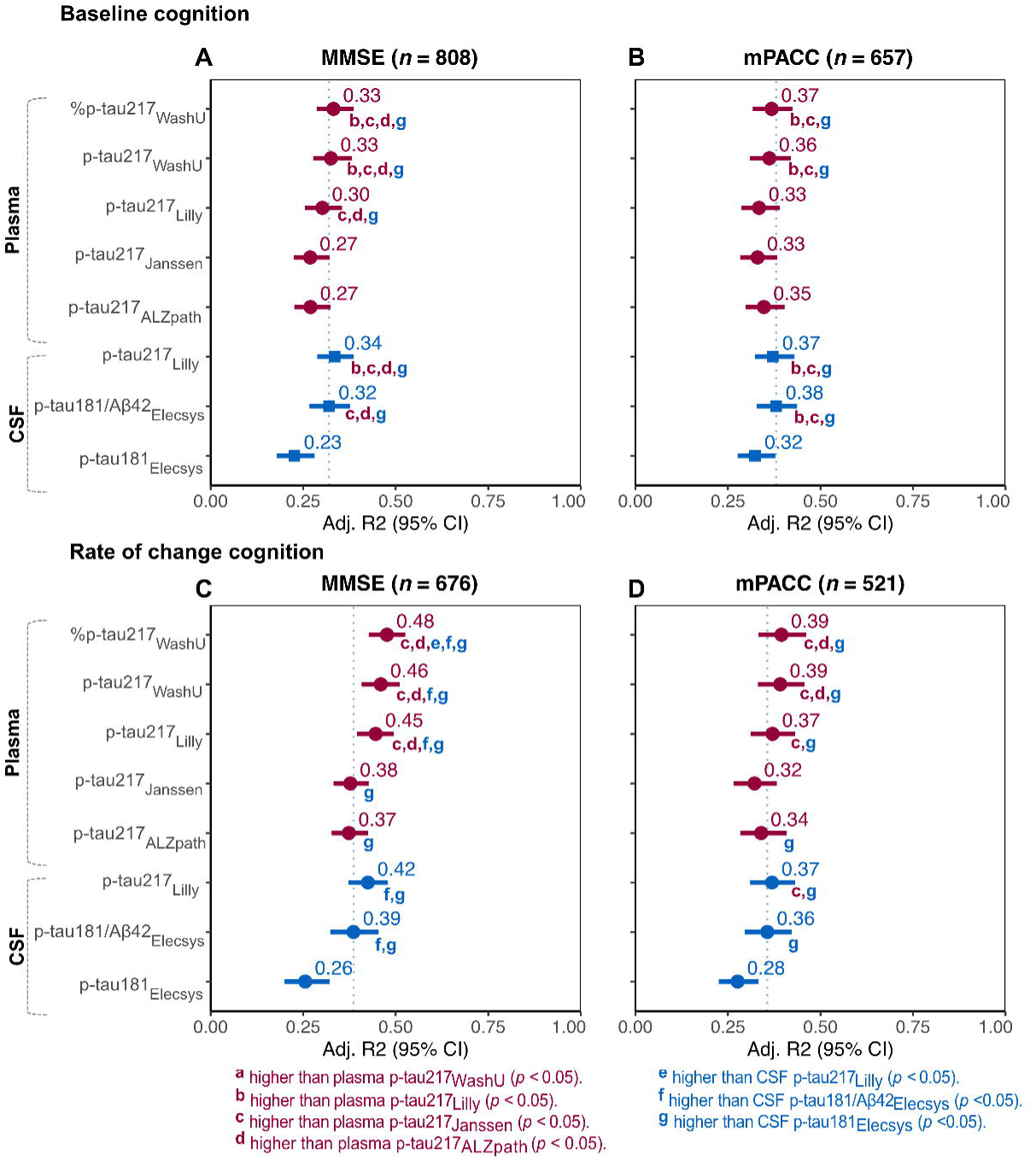
R^2^ comparisons with cognition as outcome. Squares represent the AUC or accuracy, and bars represent 95%CI. The dashed line is drawn at CSF p-tau181/Aβ42_Elecsys_, to facilitate comparing the other tests to the current approved FDA-approved test. Significant differences between assays were assessed through bootstrapping and all p-values were FDR-corrected. For the MMSE models, participants with non-AD dementia were excluded. For the mPACC models, participants with dementia were excluded. Cross-sectional: cognition ∼ biomarker + age + sex + education. Longitudinal: individual rate of change in cognition ∼ biomarker + age + sex + education. *Abbreviations*: Aβ, amyloid-beta; CI, confidence interval; FDR, false discovery rate.

We next examined the biomarker associations with cognition longitudinally. The results were generally similar for both longitudinal analyses of MMSE (**Fig. 5C**) and mPACC (**Fig. 5D**). Further information is given in **Supplementary Table 3** and **Supplementary Fig. 8 & 9**. Plasma %p-tau217_WashU_ had the strongest association with change in MMSE (R^2^: 0.48), which was significantly higher than all CSF tests (R^2^: 0.26-0.42) as well as plasma p-tau217_Janssen_ (R^2^: 0.38) and p-tau217_ALZpath_ (R^2^: 0.37, all *P*_diff_ < 0.025). Plasma p-tau217_Lilly_ (R^2^: 0.45) showed a stronger association with change in MMSE than plasma p-tau217_Janssen_ (R^2^: 0.38), plasma p-tau217_ALZpath_ (R^2^: 0.37), CSF p-tau181/Aβ42_Elecsys_ (R^2^: 0.39) and CSF p-tau181_Elecsys_ (R^2^: 0.26, all *P*_diff_ < 0.030). No differences between plasma p-tau217_Janssen_ and plasma p-tau217_ALZpath_ were observed here. For the longitudinal of change in mPACC, fewer differences between assays were observed. Plasma %p-tau217_WashU_ (R^2^: 0.40) performed better than plasma p-tau217_Janssen_ (R^2^: 0.32) and plasma p-tau217_ALZpath_ (R^2^: 0.34, all *P*_diff_ < 0.035). When comparing plasma immunoassays, plasma p-tau217_Lilly_ (R^2^: 0.37) performed better than plasma p-tau217_Janssen_ (R^2^: 0.32, *P*_diff_ = 0.018), but not better than plasma p-tau217_ALZpath_ (R^2^: 0.34). Plasma p-tau217_Janssen_ and plasma p-tau217_ALZpath_ performed similarly (*P*_diff_ = 0.210).

### Subgroups analyses in cognitively unimpaired and impaired individuals

All analyses were repeated when analyzing CU and CI individuals separately. In general, the results were similar to the ones obtained when analyzing the whole cohort, but a few differences were observed. In CI individuals, no significant differences were observed between plasma %p-tau217_WashU_, plasma p-tau217_Lilly_ and plasma p-tau217_Janssen_ regarding the rate of change in tau-PET values (R^2^ between 0.50-0.54). Further, all four plasma biomarkers performed significantly better in CI individuals than plasma p-tau217_ALZpath_ in this context (*P*_diff_ < 0.012). All results regarding the subgroup analyses can be found in **Supplementary Table 4** and **Supplementary Fig. 10-14**.

### Replication in the Knight ADRC cohort

We replicated the main cross-sectional analyses in the Knight ADRC cohort (*n* = 219) when using Aβ-PET and cognition as the outcomes and the following predictors: plasma %-ptau217_WashU_, plasma p-tau217_WashU_, plasma p-tau217_Lilly_, CSF p-tau181/Aβ42_Lumipulse_ and CSF p-tau181_Lumipulse_ (for demographics see **Supplementary Table 5**). In this cohort, plasma %p-tau217_WashU_ had a significantly higher AUC (AUC: 0.97) than plasma p-tau217_Lilly_ (AUC: 0.93, *P*_diff_ = 0.020) for detecting Aβ-PET positive participants. Plasma %p-tau217_WashU_ (AUC: 0.97) also performed significantly better than CSF p-tau181/Aβ_Lumipulse_ (AUC: 0.87, *P*_diff_ = 0.001) (**Fig. 6**). When examining associations with baseline Aβ-PET values, plasma %p-tau217_WashU_ showed a significantly better association (R^2^: 0.61) than plasma p-tau217_WashU_ (R^2^: 0.54), plasma p-tau217_Lilly_ (R^2^: 0.46) and CSF p-tau181/Aβ_Lumipulse_ (R^2^: 0.42, all *P*_diff_ < 0.004). No differences were observed between biomarkers regarding their associations with cognition.

**Figure 6.**
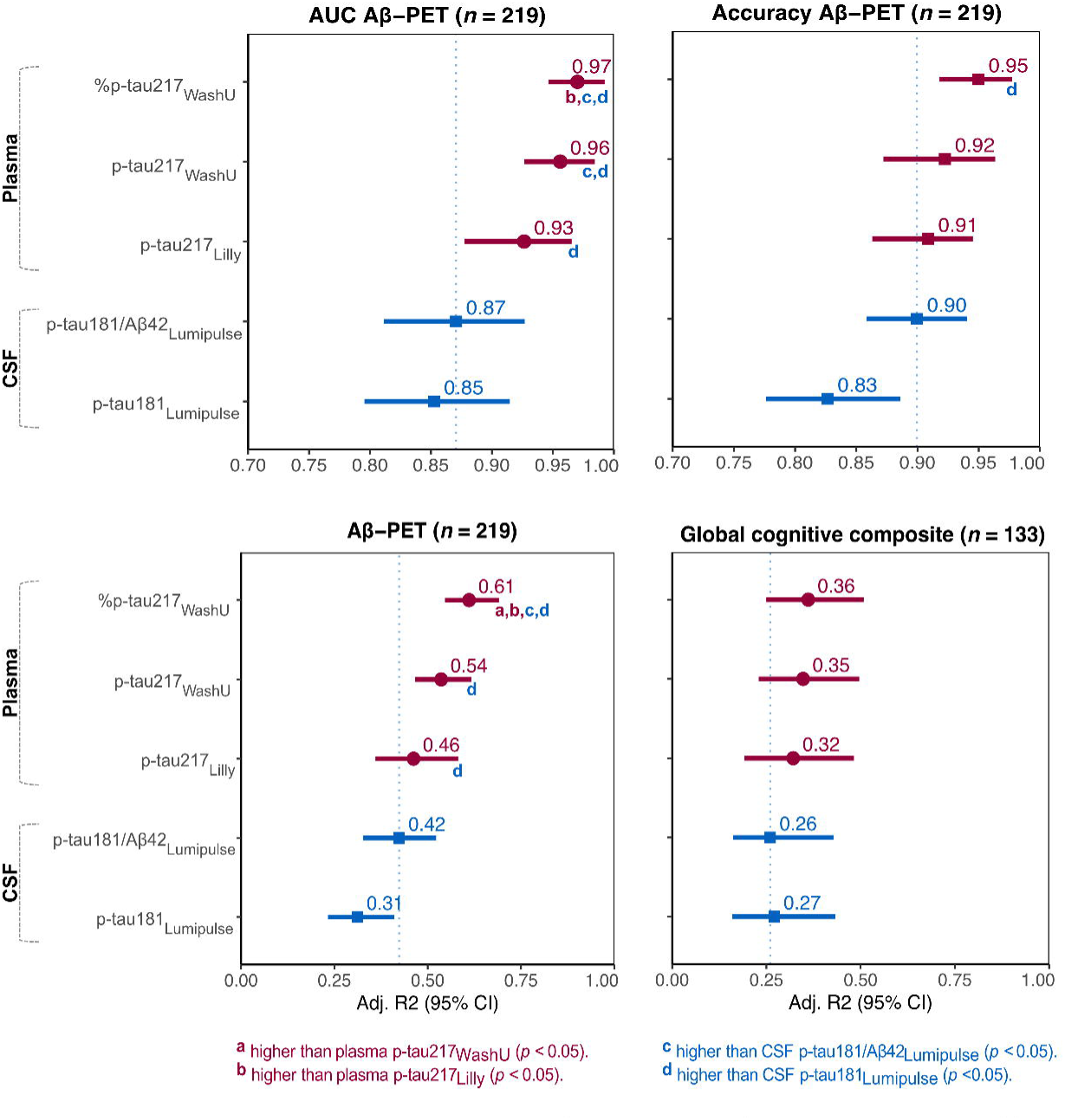
Replication in the Knight ADRC cohort. Dots and squares represent the AUC, accuracy or R2, and bars represent 95%CI. The dashed line is drawn at CSF p-tau181/Aβ42_Lumipulse_, to facilitate comparing the other tests to the current approved FDA-approved test. The global cognitive composite was a composite of several cognitive tests, z-scored with CU Aβ-individuals as reference group. Significant differences between assays were assessed through bootstrapping and all p-values were FDR-corrected. Linear model Aβ-PET: Aβ-PET ∼ biomarker + age + sex. Linear model global cognitive composite: cognition ∼ biomarker + age + sex + education. *Abbreviations*: Aβ, amyloid-beta; CI, confidence interval; CU, cognitively unimpaired; FDR, false discovery rate.

### NULISA sub-sample

In the NULISA BioFINDER-2 sub-sample (*n* = 463, **Supplementary Table 6**), we repeated the cross-sectional analyses and validated them in the Knight ADRC cohort NULISA sub-sample (*n* = 97) with the biomarkers that were available in both cohorts. Plasma %p-tau217_WashU_ outperformed plasma p-tau217_NULISA_ regarding Aβ-PET status (AUC_%WashU_: 0.96; AUC_NULISA_: 0.93, *P*_diff_ = 0.038) and Aβ-PET load (R^2^ : 0.70; R^2^ : 0.60, *P*_diff_ = 0.002) (**Supplementary Fig. 15**). Plasma %p-tau217_WashU_ was also superior to plasma p-tau217_NULISA_ regarding tau-PET status (AUC_%WashU_: 0.96; AUC_NULISA_: 0.93, *P*_diff_ = 0.038), and tau-PET values (R^2^ : 0.43; R^2^ : 0.34, *P*_diff_ = 0.048). In the Knight ADRC cohort, no differences were observed regarding Aβ-PET status, but plasma %p-tau217_WashU_ showed a significantly larger associations with Aβ-PET values than plasma p-tau217_NULISA_ (R^2^ : 0.62; R^2^ : 0.31, *P*_diff_ = 0.001) (**Supplementary Fig. 16**).

## Discussion

In this study, we performed a head-to-head comparison of different key plasma p-tau217 tests, including both MS- and immunoassay-based methods. We compared blood and CSF tests to determine Aβ- and tau-PET status at baseline, and how they were associated with baseline levels and the rate of change in Aβ- and tau-PET values and cognitive test scores. All plasma p-tau217 tests showed high AUCs and accuracies when identifying individuals with an abnormal Aβ- and tau-PET status. Across the different analyses, plasma %p-tau217_WashU_ (measured with MS) performed consistently superior to all the immunoassay tests. Further, plasma %p-tau217_WashU_ performance was non-inferior, or even superior to FDA-approved CSF measures across all analyses, as previously shown for Aβ- and tau-PET status^7^, but we now also show similar findings for either PET load or cognitive function cross-sectionally as well as longitudinally. Further, we found that among the tested immunoassays, plasma p-tau217_Lilly_ often performed better than plasma p-tau217_Janssen_ and was non-inferior to the CSF tests in most models. Plasma p-tau217_Janssen_ performed similarly to p-tau217_ALZpath_ when using tau-PET as the outcome, but p-tau217_ALZpath_ often performed better when using Aβ-PET as the outcome. CSF p-tau181, which is typically used in a clinical setting for work-up of AD patients, however, was in almost all cases significantly inferior to the other biomarkers.

One of the main questions concerning the plasma assays is the distinction between MS and immuno-based approaches. Whereas MS methods allow simultaneous quantification of different peptides, but they often come at a higher cost due to more expensive equipment and lower analysis throughput.^41^ Immunoassays, on the other hand, provide a cheaper alternative, although their results may be subject to higher measurement variability.^41,42^ Previously, plasma %p-tau217_WashU_ has demonstrated superior performance compared to immunoassays^7,8^. We have shown in a different cohort that plasma p-tau217_WashU_ had a very high accuracy for discriminating between a normal and abnormal CSF Aβ status in 135 MCI patients and had the highest AUC to identify patients who progressed to dementia, performing significantly better than immunoassays^8^. In the current study, we replicated and extended these results with additional p-tau217 tests in a larger sample consisting of both cognitively unimpaired and cognitively impaired individuals. When discriminating between normal and abnormal Aβ-PET, no significant differences were observed between MS plasma p-tau217_WashU_ (not the ratio) and plasma p-tau217_Lilly_ in the current study, which was replicated in the validation cohort. Thus, the significant differences in performance observed here may be due to implementation of the ratio phosphorylated to non-phosphorylated rather than measurement technique. For tau-PET load, however, both MS-based plasma %p-tau217_WashU_ and plasma p-tau217_WashU_ performed significantly better than immunoassay-based plasma p-tau217_Lilly_ and plasma p-tau217_Janssen_, suggesting MS-based methods may be more precise than immunoassays, at least those included in this study, in identifying patients at later stages of the disease.

However, there are also notable differences between the different immunoassays. For instance, the immunoassays included in this head-to-head comparison did not always showcase equal performance. Although all immunoassays performed very well, plasma p-tau217_Lilly_ and plasma p-tau217_ALZpath both_ had a significantly higher AUC for Aβ-PET status and showed stronger associations with baseline and longitudinal Aβ-PET load than plasma p-tau217_Janssen_. Plasma p-tau217_Lilly_ additionally showed significantly stronger associations with tau-PET load, as well as with the longitudinal accumulation of both Aβ- and tau pathology, and was more strongly associated with the rate of change in cognition, than both plasma p-tau217_Janssen_ and plasma p-tau217_ALZpath_. A recent study comparing plasma p-tau217_Janssen_ to p-tau217_ALZpath_ found that both assays were highly associated with Aβ- and tau-PET SUVR values as well as PET status in a sample (*n* = 294) consisting of CU and CI individuals.^13^ Another previous study in a different cohort of ours (*n* = 147) compared plasma p-tau217_Lilly_ to plasma p-tau217_Janssen_ in MCI patients and found they were similarly associated with Aβ-status in CSF and annual change in MMSE scores.^12^ Although plasma p-tau217_Janssen_ performed similarly to other assays in these studies, we found that when comparing plasma p-tau217_Janssen_ to other assays, its performance is slightly, but significantly, worse in its associations with Aβ-PET. The current study population consists of a larger sample, which increases the power to detect more subtle differences in assay performances. These differences may be explained by the differences in assays; the p-tau217_Janssen_ assay additionally targets p-tau212.^27^ This cross-reactivity might contribute to its overall slightly inferior performance compared to plasma p-tau217_Lilly_. Another difference between the immunoassays is that the detection antibody used in the Lilly assay does not detect big tau, which is mainly released from peripheral sources, unlike the other immunoassays used in the present study. Noteworthy, in all analyses, CSF p-tau181_Elecsys_ showed the worst performance out of all fluid biomarker tests. On the other hand, the plasma p-tau217 measures, as well as CSF p-tau217_Lilly_, showed stronger associations with disease pathology and cognition, suggesting the superiority of p-tau217 in plasma as well as in CSF over p-tau181 in CSF. This is in line with previous results^24,43^ and may be important for clinical use of fluid biomarkers.

Recently, the Global CEO Initiative on Alzheimer’s disease defined acceptable performance of blood biomarker tests of Aβ pathology.^15^ They suggested that a test should have a sensitivity and specificity of at least 90% to be used as a stand-alone confirmatory test where a subsequent test is not needed confirm to Aβ-status. Further, the recent update of the criteria for diagnosis and staging of Alzheimer’s disease lead by the Alzheimer’s Association suggested that a stand-alone biomarker test should exhibit an accuracy of 90%.^44^ In the current study, only %ptau217_WashU_ fulfilled these requirements when detecting Aβ-PET status. This was also the case when analyzing CU and CI separately. If the current results are confirmed in other large-scale cohorts, it suggests that mainly %p-tau217_WashU_ can be used as a confirmatory test. The p-tau217 immunoassays might instead be used as triaging tests where a positive plasma test result must be confirmed by a second test such as Aβ-PET. Alternatively, a two-cut point approach can be used for the p-tau217 immunoassays where test results below a lower cut point or above a higher cut point are regarded as confirmatory of a negative or positive Aβ-status, respectively. However, patients with intermediate results, which are between the two cut points, need to undergo a subsequent more high-performing test.^33^

This study has important strengths over previous studies. First, it included a large sample size consisting of both CU and CI individuals. Furthermore, we conducted a comprehensive comparison of multiple different key p-tau217 assays, including both MS- and immuno-based assays, and evaluated their performance against several and clinically relevant outcomes, with highly consistent results. We studied how the different p-tau217 tests were associated with baseline and the rate of change in Aβ- and tau-PET values, and cognitive test scores, which has not been done before. Nonetheless, this work is not without limitations. Firstly, Aβ-PET was not available in patients with a dementia diagnosis by study design, hence this patient group was excluded from analyses using Aβ-PET as an outcome. The sample was further reduced when examining longitudinal changes in AD pathology, although it still consisted of a considerably large sample. However, considering disease-modifying therapies will likely target individuals at early stages of the disease, it is important to assess how plasma p-tau217 performs as a biomarker also in cognitively unimpaired individuals alone, as we have done in the present study. Further replication of these findings in other cohorts are warranted as performance could be affected by the prevalence of amyloid positivity in the cohort, the relative distribution of participants at different stages of amyloid deposition, or the prevalence of participants with other neurological disorders, as suggested by the somewhat better performance of the MS assay in the Knight ADRC cohort. Such replications will be required to fully comprehend how blood-based biomarkers perform among different populations. ^45–47^ Biomarker performance and their accessibility and feasibility outside the research setting should additionally be further investigated. Here, it will be important to examine the additional plasma p-tau217 assays (from other vendors) that might become commercially available in the future.

In summary, all plasma p-tau217 biomarkers in this study consistently and reliably detected abnormalities in Aβ-PET and tau-PET. However, associations with Aβ-PET and tau-PET values, and with cognition, differed between the different plasma and CSF tests. Overall, plasma %p-tau217 measured with MS was superior to the immunoassay-based plasma p-tau217 tests. We also observed differences in performance between immunoassays, in which p-tau217_Lilly_ was superior to p-tau217_Janssen_ and p-tau217_ALZpath_, but the differences were in general rather minor. If the current results are replicated in other larges-cale cohorts, plasma %p-tau217 might potentially be the only blood tests that currently fulfils the suggested criteria to be used as a stand-alone confirmatory test of presence of AD pathology. However, the evaluated p-tau217 immunoassays might be more suitable for use as triaging tests where positive or intermediate test results must be confirmed with a second test like PET.

## Supplementary material

Supplementary material is available at *Brain* online.

