## Supplementary materials for "A Comprehensive Head-to-Head Comparison of Key Plasma Phosphorylated Tau 217 Biomarker Tests"

### Supplementary material

#### Plasma and CSF p-tau analysis

***P-tau-MS$_{WashU}$***

P-tau217 concentrations and non-phosphorylated tau217 occupancies were measured at the Department of Neurology, Washington University School of Medicine (St. Louis, MO, USA) with the multiplex mass spectrometry assay developed at Washington University (MS-WashU). The tau antibody was immobilized on magnetic beads for immunopurification, followed by solid phase extraction at the protein level and extraction at the peptide level. 15N-labeled tau was added to one ml of plasma which was then immuno-precipitated with the Tau1 antibody (anti tau-192-199) crosslinked to Dynabeads M270 Epoxy magnetic beads. The tau precipitate was eluted and then extracted on a HLB μelution plate. A mixture of labeled synthetic peptides was added for each analyte after drying the eluate. The sample was then digested with trypsin, which was extracted on a HLB μ-elution plate; the eluate was dried then resuspended prior to nanoLC-MS/HRMS analysis. The intra-assay and inter-assay variability were assessed by means of a replicate analyses of a plasma pool from normal controls with a normal tau217 status[1].

***P-tau$_{Lilly}$***

P-tau217 was measured at the Clinical Memory Research Unit, Lund University (Sweden) by means of the Meso Scale Discovery (MSD) immunoassay developed by Lilly Research Laboratories[2-5]. The plasma samples were analyzed in duplicates as recommended in the published protocol. Biotinylated-IBA493 (p-tau217) was used as the capture antibody and SULFO-TAG-4G10-E2 as the detector. Calibration took place by means of a synthetic p-tau217 peptide.

***P-tau217$_{Janssen}$***

P-tau217 was measured at Janssen (La Jolla, CA, USA), with a Simoa assay developed by Johnson and Johnson Innovative Medicine[6-8]. PT3 was used as a capture antibody whose binding requires phosphorylation at aa217 and is enhanced from additional phosphorylation at aa212. HT43 (anti-tau) was used as the detector. Calibration took place by means of a synthetic p-tau212/217 peptide.

***P-tau217$_{ALZpath}$***

The ALZpath pTau217 p-tau217 assay uses a proprietary monoclonal p-tau217 specific capture antiboy, an N-terminal detecting antibody, and a peptide calibrator[9]. The assay has been confirmed to be fit-for-purpose, demonstrating a detection limit between 0.0052 and 0.0074 pg/mL, with a functional quantification lower limit of 0.06 pg/mL, and a dynamic range spanning from 0.007 to 30 pg/mL. The assay's spike recovery rate for the endogenous analyte was 80%, with intrarun precision ranging from 0.5% to 13% and interrun precision ranging from 9.2% to 15.7%.

***P-tau217$_{NULISA}$***

Nucleic Acid Linked Immuno-Sandwich Assay (NULISA™) Measurements (NULISA™) is a new technology that employs a dual capture and to the attomole level. The process involves DNA-conjugated antibodies forming immunocomplexes with the target protein in solution. These immunocomplexes are then sequentially purified: first using oligo-dT beads that bind to the polyA sequence attached to the capture antibody, and then with streptavidin beads that bind to the biotin on the detection antibody. Subsequently, a DNA reporter is generated through proximity ligation and amplified by PCR to produce next-generation sequencing libraries. The DNA oligonucleotide includes a barcode to facilitate the de-multiplexing of targeted proteins and sample identification. This technology can measure over 200 proteins in a mere 10µL sample. Concentrations are reported in NULISA Protein Quantification (NPQ) units, which normalize the sequence quantification counts by accounting for intraplate and intensity variability and then log2-transforming the data to approximate normality[10].

### Supplementary Tables

**Supplementary Table 1. AUC and accuracy, full sample (*n* = 998).** Significant differences between assays were assessed through bootstrapping. In brief, a bootstrap distribution of the differences in AUC and accuracy of two different biomarkers was generated (n = 1000 iterations), and their 95% CI. Then, p-values were derived from the distributed differences in values, which were subsequently FDR-corrected. *Abbreviations*: Aβ, amyloid-beta; CI, confidence interval; FDR, false discovery rate.

| | | **Aβ-PET** | | **Tau-PET** | |
|---|---|---|---|---|---|
| | | **AUC (95% CI)** | **Accuracy (95% CI)** | **AUC (95% CI)** | **Accuracy (95% CI)** |
| Plasma | %p-tau217$_{WashU}$ | 0.963 (0.946,0.978) | 0.931 (0.909,0.951) | 0.970 (0.960,0.978) | 0.877 (0.866,0.923) |
| | p-tau217$_{WashU}$ | 0.946 (0.926,0.963) | 0.885 (0.867,0.922) | 0.964 (0.953,0.973) | 0.889 (0.845,0.922) |
| | p-tau217$_{Lilly}$ | 0.939 (0.919,0.956) | 0.883 (0.854,0.908) | 0.956 (0.944,0.968) | 0.877 (0.841,0.915) |
| | p-tau217$_{Janssen}$ | 0.907 (0.883,0.930) | 0.844 (0.814,0.878) | 0.949 (0.936,0.962) | 0.849 (0.839,0.904) |
| | p-tau217$_{ALZpath}$ | 0.930 (0.910,0.949) | 0.873 (0.846,0.905) | 0.943 (0.929,0.956) | 0.873 (0.822,0.901) |
| CSF | p-tau217$_{Lilly}$ | 0.949 (0.929,0.968) | 0.899 (0.873,0.928) | 0.963 (0.953,0.974) | 0.887 (0.855,0.918) |
| | p-tau181/Aβ42$_{Elecsys}$ | 0.939 (0.919,0.960) | 0.892 (0.873,0.927) | 0.941 (0.925,0.956) | 0.873 (0.825,0.899) |
| | p-tau181$_{Elecsys}$ | 0.846 (0.813,0.879) | 0.805 (0.748,0.839) | 0.873 (0.845,0.900) | 0.797 (0.741,0.846) |

**Supplementary Table 2. Outcomes of cross-sectional linear regression models, full sample (*n* = 998).** Linear regression models were conducted with biomarkers as predictors and age and sex as covariates. Education was included as an additional covariate in the cognition models. For the MMSE models, participants with non-AD dementia were excluded. mPACC models were restricted to CU and MCI participants. Significant differences between assays were assessed through bootstrapping. In brief, a bootstrap distribution of the difference in $R^2$ of two different biomarkers was generated (n = 1000 iterations). Then, p-values were derived from the distributed differences in $R^2$ values, which were subsequently FDR-corrected. Reported beta's are standardized. Linear regression: PET SUVR or cognition ~ biomarker + age + sex (+ education). *Abbreviations*: Aβ, amyloid-beta; CI, confidence interval; FDR, false discovery rate.

| | **Aβ-PET** *n* **= 694** | | **Tau-PET** *n* **= 961** | | **MMSE** *n* **= 808** | | **mPACC** *n* **= 657** | |
|---|---|---|---|---|---|---|---|---|
| | $\beta_{std}$ (SE) | $R^2$ (95% CI) | $\beta_{std}$ (SE) | $R^2$ (95% CI) | $\beta_{std}$ (SE) | $R^2$ (95% CI) | $\beta_{std}$ (SE) | $R^2$ (95% CI) |
| **Plasma** | | | | | | | | |
| %p-tau217$_{WashU}$ | 0.95 (0.02) | 0.72 (0.68,0.76) | 0.73 (0.02) | 0.51 (0.47,0.55) | -0.53 (0.03) | 0.33 (0.29,0.39) | -0.37 (0.04) | 0.37 (0.32,0.43) |
| p-tau217$_{WashU}$ | 0.91 (0.03) | 0.65 (0.60,0.69) | 0.72 (0.02) | 0.49 (0.45,0.53) | -0.52 (0.03) | 0.33 (0.28,0.38) | -0.37 (0.04) | 0.36 (0.31,0.42) |
| p-tau217$_{Lilly}$ | 0.81 (0.03) | 0.58 (0.54,0.63) | 0.66 (0.02) | 0.45 (0.40,0.49) | -0.48 (0.03) | 0.30 (0.26,0.36) | -0.28 (0.04) | 0.33 (0.29,0.39) |
| p-tau217$_{Janssen}$ | 0.69 (0.03) | 0.47 (0.42,0.52) | 0.63 (0.03) | 0.39 (0.34,0.44) | -0.45 (0.03) | 0.27 (0.23,0.32) | -0.26 (0.04) | 0.33 (0.28,0.38) |
| p-tau217$_{ALZpath}$ | 0.81 (0.03) | 0.57 (0.52,0.61) | 0.64 (0.03) | 0.38 (0.34,0.43) | -0.47 (0.03) | 0.27 (0.23,0.32) | -0.32 (0.04) | 0.35 (0.30,0.40) |
| **CSF** | | | | | | | | |
| p-tau217$_{Lilly}$ | 0.88 (0.03) | 0.65 (0.61,0.69) | 0.71 (0.03) | 0.47 (0.43,0.51) | -0.53 (0.03) | 0.34 (0.29,0.39) | -0.37 (0.04) | 0.37 (0.32,0.43) |
| p-tau181/Aβ42$_{Elecsys}$ | 0.87 (0.03) | 0.62 (0.58,0.67) | 0.70 (0.03) | 0.44 (0.39,0.49) | -0.52 (0.03) | 0.32 (0.27,0.38) | -0.39 (0.04) | 0.38 (0.33,0.44) |
| p-tau181$_{Elecsys}$ | 0.60 (0.04) | 0.38 (0.32,0.44) | 0.55 (0.03) | 0.29 (0.24,0.34) | -0.41 (0.03) | 0.23 (0.18,0.28) | -0.25 (0.04) | 0.32 (0.28,0.38) |

**Supplementary Table 3. Outcomes of longitudinal $R^2$ models, full sample (*n* = 998).** Linear mixed models were conducted with time-between-visits as predictors to obtain the rate of change. Then, linear regression models were carried out with biomarkers as predictors and age and sex as covariates. Education was included as an additional covariate in the cognition linear regression models. For the MMSE models, participants with non-AD dementia were excluded. mPACC models were restricted to CU and MCI participants. Significant differences between assays were then assessed through bootstrapping. In brief, a bootstrap distribution of the difference in $R^2$ of two different biomarkers was generated (n = 1000 iterations). Then, p-values were derived from the distributed differences in $R^2$ values, which were subsequently FDR-corrected. Linear mixed model: PET SUVR or cognition ~ time between visits. Linear regression: rate of change in PET SUVR or cognition ~ biomarker + age + sex (+ education). *Abbreviations*: Aβ, amyloid-beta; CI, confidence interval; FDR, false discovery rate.

| | **Aβ-PET** *n* **= 448** | **Tau-PET** *n* **= 524** | **MMSE** *n* **= 676** | **mPACC** *n* **= 521** |
|---|---|---|---|---|
| | **$R^2$ (95% CI)** | **$R^2$ (95% CI)** | **$R^2$ (95% CI)** | **$R^2$ (95% CI)** |
| **Plasma** | | | | |
| $\%$p-tau217$_{WashU}$ | 0.53 (0.47,0.59) | 0.50 (0.44,0.56) | 0.48 (0.43,0.53) | 0.40 (0.33,0.46) |
| p-tau217$_{WashU}$ | 0.48 (0.42,0.54) | 0.48 (0.42,0.54) | 0.46 (0.41,0.51) | 0.39 (0.33,0.46) |
| p-tau217$_{Lilly}$ | 0.38 (0.32,0.45) | 0.43 (0.36,0.50) | 0.45 (0.40,0.50) | 0.37 (0.31,0.43) |
| p-tau217$_{Janssen}$ | 0.31 (0.25,0.38) | 0.36 (0.29,0.43) | 0.38 (0.33,0.43) | 0.32 (0.27,0.38) |
| p-tau217$_{ALZpath}$ | 0.37 (0.31,0.44) | 0.35 (0.29,0.42) | 0.37 (0.33,0.43) | 0.34 (0.28,0.41) |
| CSF | | | | |
| p-tau217$_{Lilly}$ | 0.46 (0.41,0.53) | 0.44 (0.38,0.51) | 0.43 (0.37,0.48) | 0.37 (0.31,0.43) |
| p-tau181/Aβ42$_{Elecsys}$ | 0.48 (0.42,0.54) | 0.41 (0.34,0.49) | 0.39 (0.32,0.45) | 0.36 (0.30,0.42) |
| p-tau181$_{Elecsys}$ | 0.25 (0.19,0.32) | 0.26 (0.20,0.33) | 0.26 (0.20,0.32) | 0.28 (0.23,0.33) |

**Supplementary Table 4. Identification of abnormal Aβ- or tau-PET status in CU vs. CI individuals.** Sensitivity, specificity, positive predictive values (PPV) and negative predictive values (NPV) were calculated using the threshold derived from the optimization of the Youden index ('pROC' package). [a]Aβ-PET cutoff: ≥1.033. Participants with dementia do not undergo Aβ-PET by study design (missing $n$ = 37 in CU, $n$ = 267 in CI). [b]Tau-PET cutoff: ≥1.362. Tau-PET is missing for $n$ = 5 in CU, $n$ = 18 in CI. Abbreviations: Aβ, amyloid-beta; CI, cognitively impaired; CU, cognitively unimpaired.

| | | **CU ($n$ = 514)** | | | | **CI ($n$ = 484)** | | | |
|---|---|---|---|---|---|---|---|---|---|
| **Aβ-PET[a]** | | *Sensitivity* | *Specificity* | *PPV* | *NPV* | *Sensitivity* | *Specificity* | *PPV* | *NPV* |
| Plasma | %p-tau217$_{WashU}$ | 0.907 | 0.943 | 0.822 | 0.972 | 0.912 | 0.950 | 0.969 | 0.864 |
| | p-tau217$_{WashU}$ | 0.925 | 0.870 | 0.673 | 0.976 | 0.912 | 0.887 | 0.933 | 0.855 |
| | p-tau217$_{Lilly}$ | 0.832 | 0.895 | 0.695 | 0.948 | 0.898 | 0.863 | 0.918 | 0.831 |
| | p-tau217$_{Janssen}$ | 0.813 | 0.843 | 0.600 | 0.94 | 0.905 | 0.825 | 0.899 | 0.835 |
| | p-tau217$_{ALZpath}$ | 0.888 | 0.824 | 0.594 | 0.962 | 0.883 | 0.875 | 0.924 | 0.814 |
| CSF | p-tau217$_{Lilly}$ | 0.879 | 0.919 | 0.758 | 0.963 | 0.818 | 0.975 | 0.982 | 0.757 |
| | p-tau181/Aβ42$_{Elecsys}$ | 0.860 | 0.905 | 0.724 | 0.957 | 0.832 | 0.938 | 0.958 | 0.765 |
| | p-tau181$_{Elecsys}$ | 0.757 | 0.800 | 0.523 | 0.919 | 0.642 | 0.938 | 0.946 | 0.605 |
| **Tau-PET[b]** | | | | | | | | | |
| Plasma | %p-tau217$_{WashU}$ | 1 | 0.867 | 0.256 | 1 | 0.927 | 0.870 | 0.822 | 0.949 |
| | p-tau217$_{WashU}$ | 1 | 0.853 | 0.237 | 1 | 0.849 | 0.906 | 0.854 | 0.903 |
| | p-tau217$_{Lilly}$ | 0.955 | 0.820 | 0.194 | 0.997 | 0.877 | 0.863 | 0.805 | 0.916 |
| | p-tau217$_{Janssen}$ | 0.864 | 0.902 | 0.288 | 0.993 | 0.927 | 0.809 | 0.758 | 0.945 |
| | p-tau217$_{ALZpath}$ | 1 | 0.84 | 0.222 | 1 | 0.816 | 0.866 | 0.798 | 0.879 |
| CSF | p-tau217$_{Lilly}$ | 1 | 0.936 | 0.415 | 1 | 0.877 | 0.874 | 0.818 | 0.917 |
| | p-tau181/Aβ42$_{Elecsys}$ | 1 | 0.902 | 0.319 | 1 | 0.922 | 0.794 | 0.743 | 0.940 |
| | p-tau181$_{Elecsys}$ | 0.955 | 0.851 | 0.226 | 0.998 | 0.726 | 0.830 | 0.734 | 0.824 |

**Supplementary Table 5. Description of sub-sample of participants with NULISA data.**

| | **Total sample** **$n$ = 463** |
|---|---|
| **Characteristics** | |
| Age (years) | 72.3 ± 7.7 (41.4 – 89.8) |
| Women, n (%) | 218 (47%) |
| Education (years) | 12.8 ± 4.0 (3 – 31) |
| MMSE score | 26.7 ± 3.6 (12 – 30) |
| mPACC score | -1.25 ± 1.44 (-6.07 – 1.49) |
| *APOE-ε4* carriers, n (%) | 193 (42%) |
| Aβ+ participants, n (%) | 244 (53%) |
| Cognitive status (Normal/SCD/MCI/Dementia) | 95 / 107 /145 / 116 |
| **PET** | |
| [$^{18}$F]flutemetamol global Aβ-PET SUVR[a] | 1.18 ± 0.31 (0.82 – 2.05) |
| [$^{18}$F]RO948 temporal-meta ROI tau-PET SUVR[a] | 1.33 ± 0.42 (0.86 – 4.29) |
| **Plasma biomarkers** | |
| %p-tau217$_{WashU}$, % | 1.80 ± 1.68 (0.24 – 12.81) |
| p-tau217$_{WashU}$ (pg/ml) | 0.00 ± 0.00 (0.00 – 0.03) |
| p-tau217$_{Lilly}$ (pg/ml) | 0.32 ± 0.29 (0.04 – 2.01) |
| p-tau217$_{NULISA}$ | 11.50 ± 0.75 (9.79 – 13.76) |

Data are presented as mean ± standard deviation (range), unless otherwise specified. Aβ positivity was defined with the CSF Aβ42/40 ratio. [a] Participants diagnosed with dementia do not undergo Aβ-PET by study design (missing n = 153). Tau-PET is missing for $n$ = 16. Abbreviations: Aβ, amyloid-beta; *APOE-ε4*, apolipoprotein E carriership (carrying at least one ε4 allele); CSF, cerebrospinal fluid; CU, cognitively unimpaired; MCI, mild cognitive impairment; MMSE, mini-mental state examination; PET, positron emission tomography; ROI, region of interest; SCD, subjective cognitive decline; SUVR, standardized uptake value ratio.

**Supplementary Table 6. Description of the Knight ADRC cohort.**

| | Total sample *n* = 219 | Subset ALZpath n = 97 |
|---|---|---|
| **Characteristics** | | |
| Age (years) | 62.1 ± 6.2 (60.0 - 91.4) | 70.6 ± 6.0 (60.1 – 88.2) |
| Women, n (%) | 118 (54%) | 58 (60%) |
| Education (years) | 16.4 ± 2.3 (10 – 20) | 16.3 ± 2.3 (12– 20) |
| Race, (% white) | 202 (92%) | 91 (94%) |
| CDR score (0 / 0.5 / 1) | 192 (88%) / 25 (11%) / 2 (1%) | 83 (86%) / 12 (12%) / 2 (2%) |
| *APOE-ε4* carriers, n (%) | 72 (33%) | 32 (33%) |
| Aβ+ participants, n (%) | 84 (38%) | 40 (41%) |
| Global cognitive composite | -0.5 ± 1.1 (-4.7 – 1.3) | -0.8 ± 1.4 (-5.3 – 1.2) |
| **PET** | | |
| Global Aβ-PET SUVR[a] | 1.33 ± 0.65 (0.59 – 3.95) | 1.33 ± 0.62 (0.69 – 3.40) |
| **Plasma biomarkers** | | |
| %p-tau217$_{WashU}$, % | 1.21 ± 1.18 (0.21 – 7.75) | 1.16 ± 1.23 (0.27 – 7.75) |
| p-tau217$_{WashU}$ (pg/ml) | 0.00 ± 0.00 (0.00 – 0.02) | 0.00 ± 0.00 (0.00 – 0.02) |
| p-tau217$_{Lilly}$ (pg/ml) | 0.29 ± 0.20 (0.08 – 1.38) | 0.29 ± 0.22 (0.09 – 1.38) |
| p-tau217$_{NULISA}$ (pg/ml) | - | 5.29 ± 3.76 (1.57 – 21.17) |
| **CSF biomarkers** | | |
| p-tau181/Aβ42$_{Lumipulse}$ | 0.00 ± 0.00 (0.00 – 0.01) | 0.00 ± 0.00 (0.00 – 0.01) |
| p-tau181$_{Lumipulse}$ (pg/ml)[6] | 1.58 ± 0.22 (1.09 – 2.34) | 1.59 ± 0.24 (1.19 – 2.34) |

Data are presented as mean ± standard deviation (range), unless otherwise specified. Aβ positivity was defined with the CSF Aβ42/40 ratio. The global cognitive composite is missing for n = 38. [a]Aβ-PET was performed with either tracer [$^{18}$F]florbetapir (AV45) or [$^{11}$C]Pittsburgh Compound B (PiB). Abbreviations: Aβ, amyloid-beta; *APOE-ε4*, apolipoprotein E carriership (carrying at least one ε4 allele); CSF, cerebrospinal fluid; CU, cognitively unimpaired; MCI, mild cognitive impairment; MMSE, mini-mental state examination; PET, positron emission tomography; ROI, region of interest; SCD, subjective cognitive decline; SUVR, standardized uptake value ratio.

**Supplementary Figure 1.**

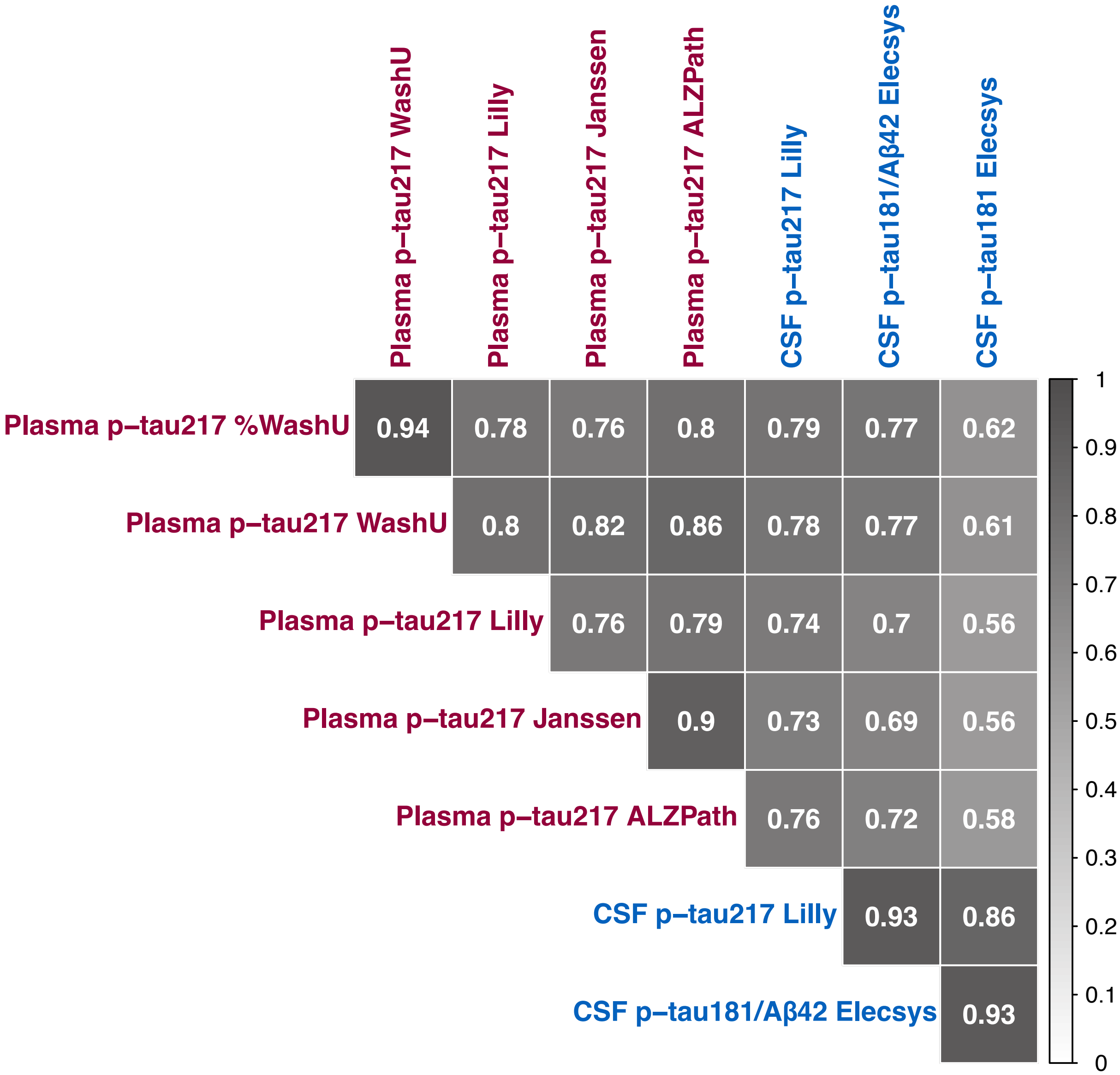


**Correlations between biomarkers across the full sample.** Heatmap showing Spearman coefficients for correlations between plasma p-tau217 assays and CSF assays ($n$ = 998).

**Supplementary Figure 2. Correlations between biomarkers and Aβ-PET.** Biomarker concentrations were z-scored based on CU, CSF Aβ− participants. Z-scores have been plotted against Aβ-PET global composite SUVRs ($n$ = 694). Lighter dots: CU Darker dots: CI. *Abbreviations*: Aβ, amyloid-beta.

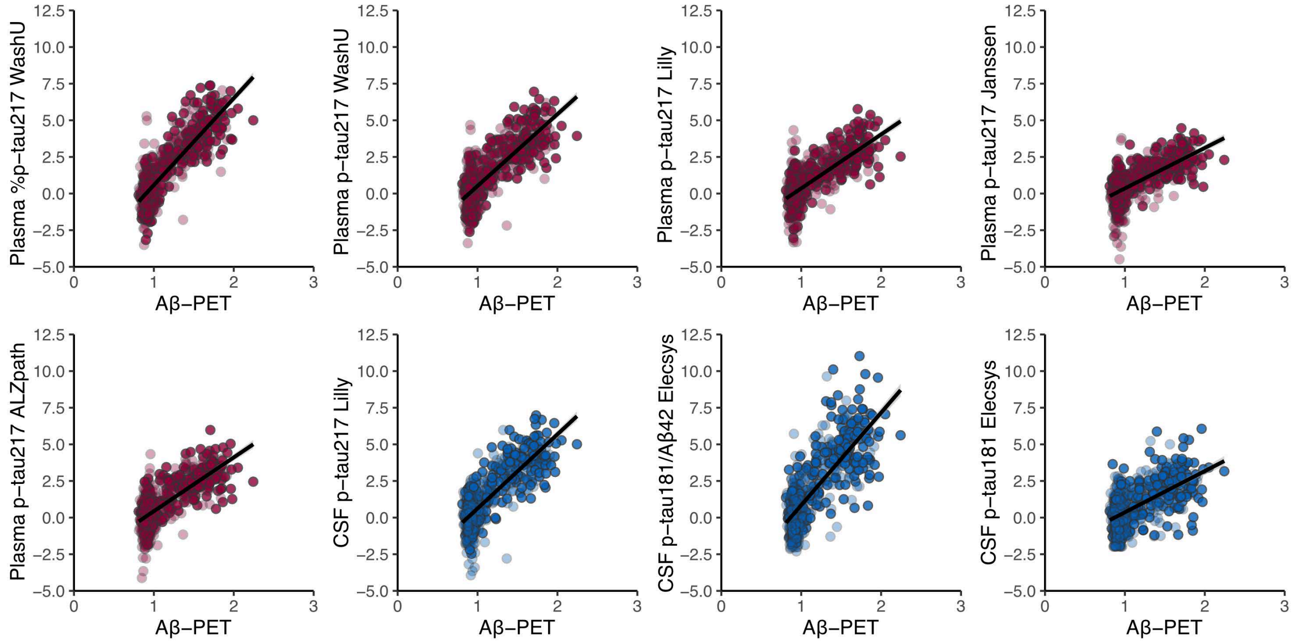


**Supplementary Figure 3. Correlations between biomarkers and the rate of change in Aβ-PET.** Biomarker concentrations were z-scored based on CU, CSF Aβ− participants. Z-scores have been plotted against the rate of change in the Aβ-PET global composite SUVRs ($n$ = 448). The rate of change was obtained from linear mixed models. Lighter dots: CU Darker dots: CI. *Abbreviations*: Aβ, amyloid-beta.

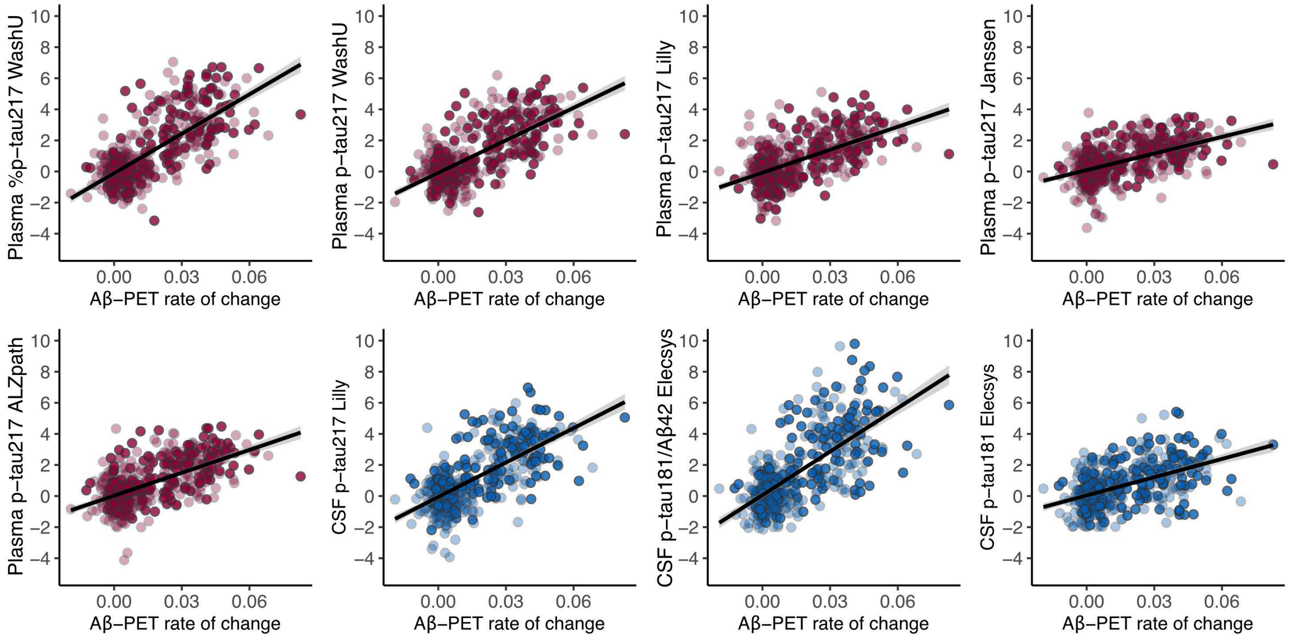

**Supplementary Figure 4. Correlations between biomarkers and tau-PET.** Biomarker concentrations were z-scored based on CU, CSF Aβ– participants. Z-scores have been plotted against tau-PET temporal-meta SUVRs (*n* = 961). Lighter dots: CU Darker dots: CI. *Abbreviations*: Aβ, amyloid-beta.

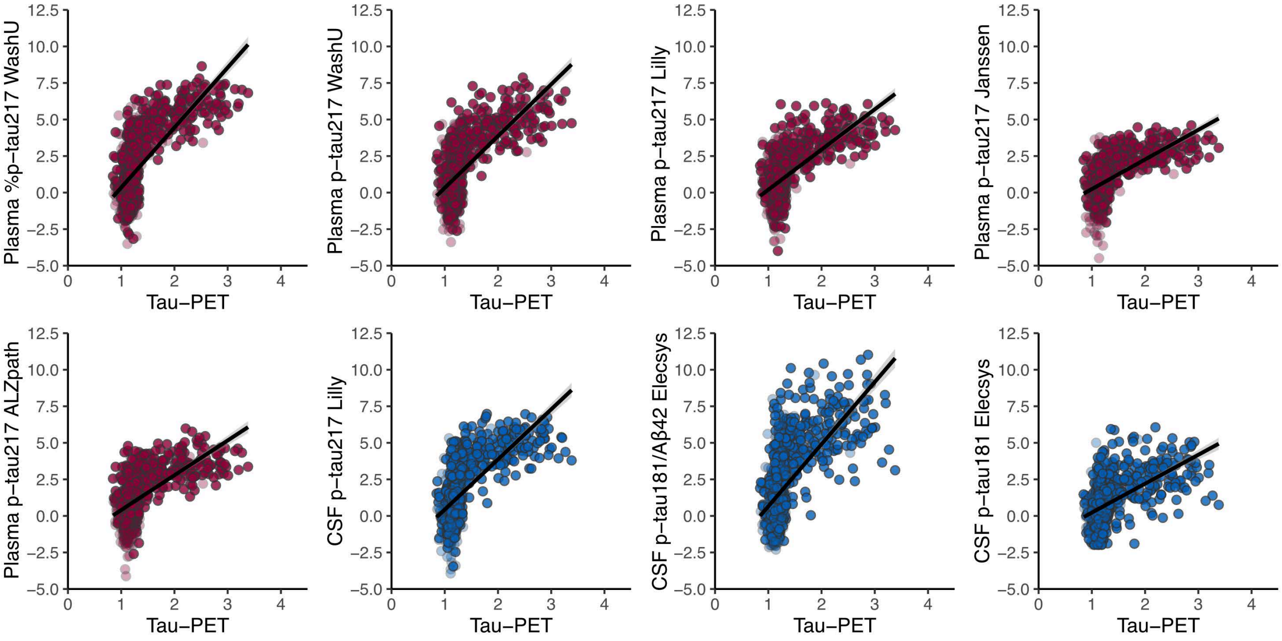


**Supplementary Figure 5. Correlations between biomarkers and the rate of change in tau-PET.** Biomarker concentrations were z-scored based on CU, CSF Aβ– participants. Z-scores have been plotted against the rate of change in tau-PET temporal-meta SUVRs (*n* = 524). The rate of change was obtained from linear mixed models. Lighter dots: CU Darker dots: CI. *Abbreviations*: Aβ, amyloid-beta.

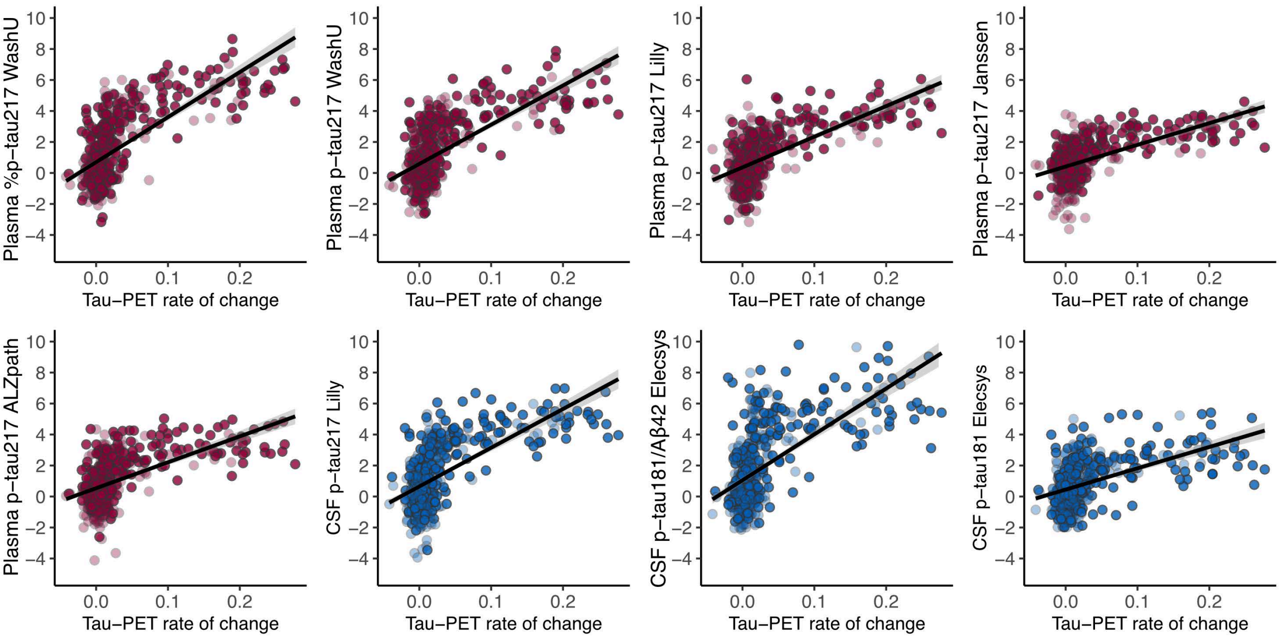

**Supplementary Figure 6. Correlations between biomarkers and MMSE scores.** Biomarker concentrations were z-scored based on CU, CSF Aβ− participants. Z-scores have been plotted against MMSE scores. Non-AD dementias were omitted ($n = 808$). Lighter dots: CU Darker dots: CI. *Abbreviations*: Aβ, amyloid-beta.

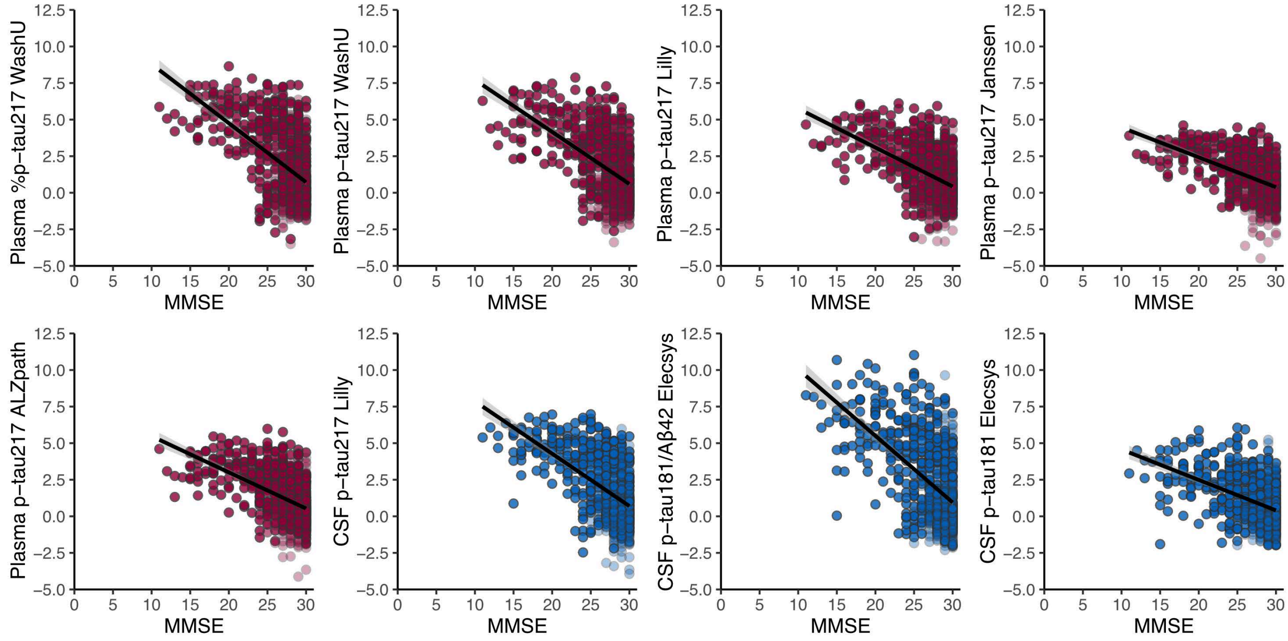


**Supplementary Figure 7. Correlations between biomarkers and mPACC scores.** Biomarker concentrations were z-scored based on CU, CSF Aβ− participants. Z-scores have been plotted against mPACC scores. Participants with dementia were omitted ($n = 657$). Lighter dots: CU Darker dots: CI. *Abbreviations*: Aβ, amyloid-beta.

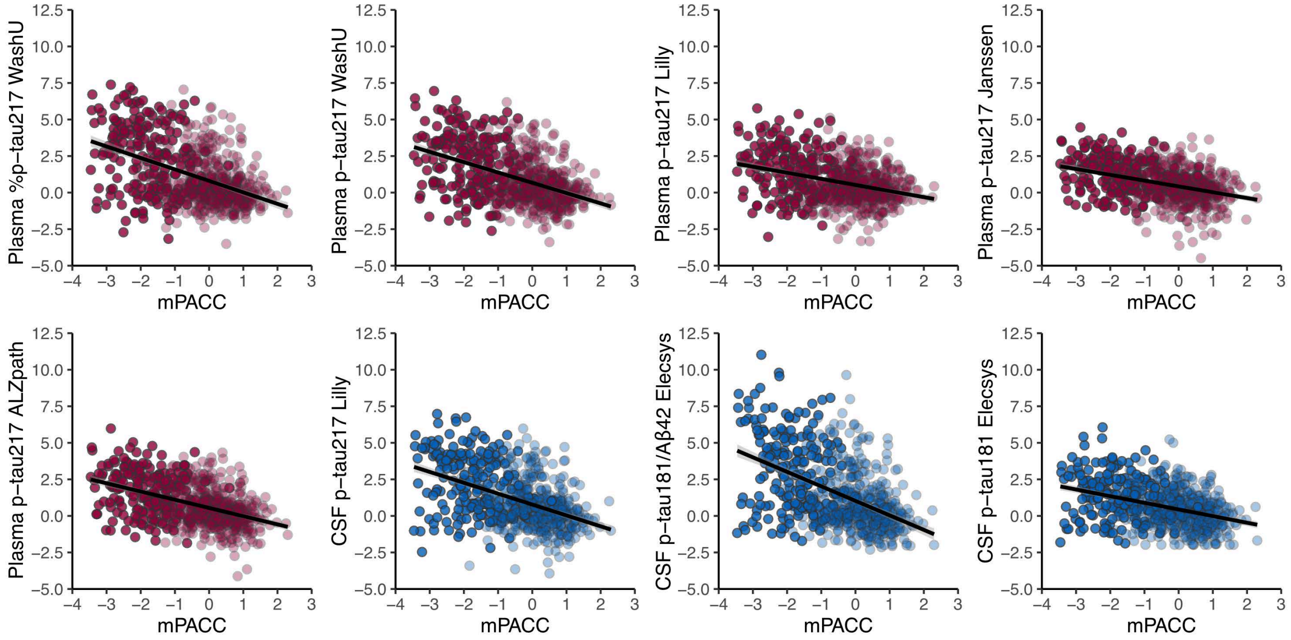

**Supplementary Figure 8. Correlations between biomarkers and the rate of change in MMSE scores.** Biomarker concentrations were z-scored based on CU, CSF Aβ− participants. Z-scores have been plotted against the rate of change in MMSE scores. Non-AD dementias were omitted (*n* = 676). The rate of change was obtained from linear mixed models. Lighter dots: CU Darker dots: CI. *Abbreviations*: Aβ, amyloid-beta.

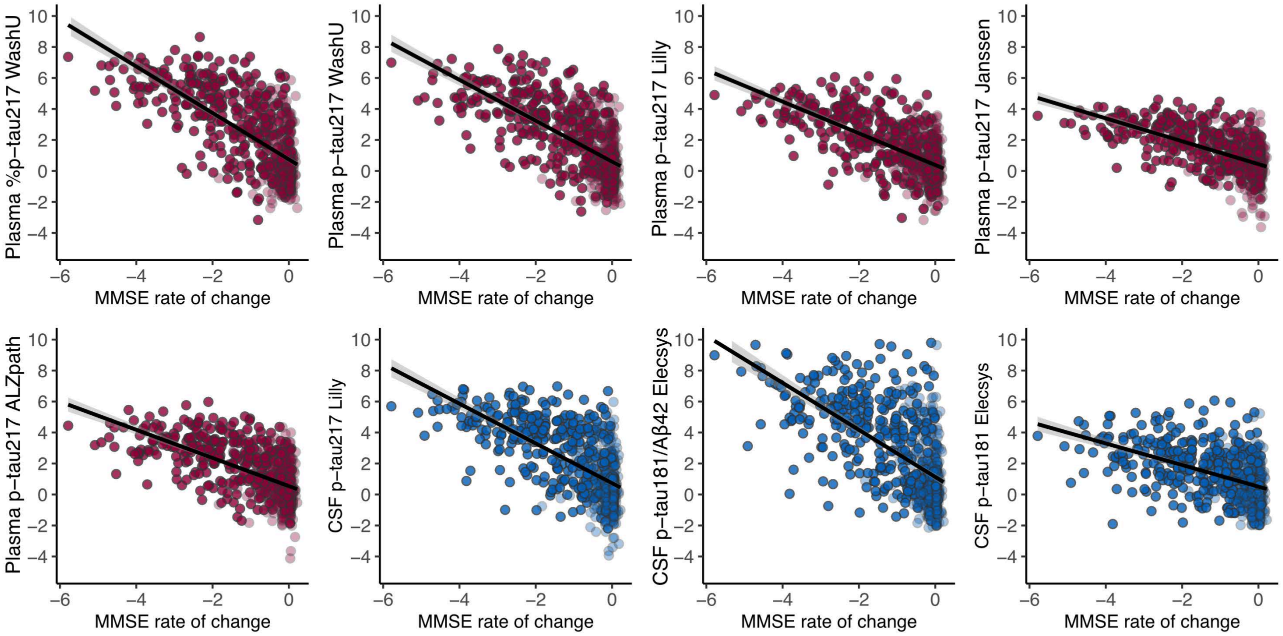


**Supplementary Figure 9. Correlations between biomarkers and the rate of change in mPACC scores.** Biomarker concentrations were z-scored based on CU, CSF Aβ− participants. Z-scores have been plotted against the rate of change in mPACC scores. Participants with dementia were omitted (*n* = 521). The rate of change was obtained from linear mixed models. Lighter dots: CU Darker dots: CI. *Abbreviations*: Aβ, amyloid-beta.

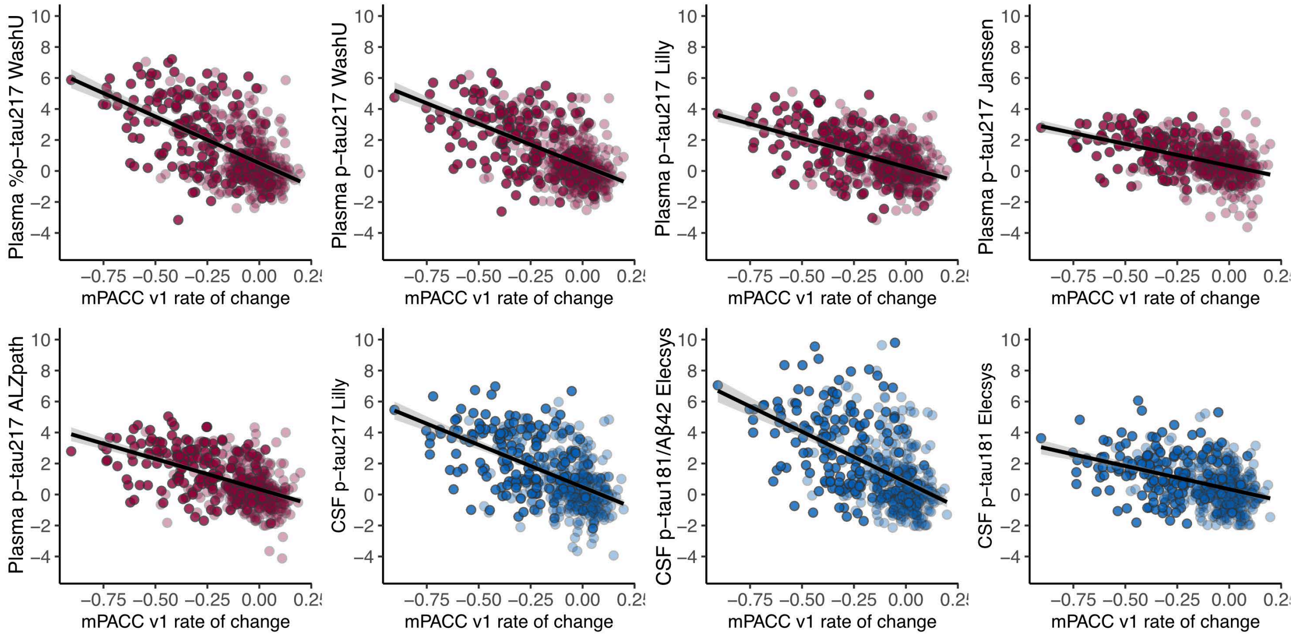

**Supplementary Figure 10. AUC comparisons in cognitively unimpaired and cognitively impaired.** The dashed line is drawn at CSF p-tau181/Aβ42$_{Elecsys}$ to facilitate comparing the other tests to the current approved FDA-approved test. Significant differences between assays were assessed through bootstrapping. In brief, a bootstrap distribution of the difference in AUC of two different biomarkers was generated (*n* = 1000 iterations), and their 95% CI. Then, p-values were derived from the distributed differences in values, which were subsequently FDR-corrected. *Abbreviations*: Aβ, amyloid-beta; CU, cognitively unimpaired; CI, cognitively impaired; 95% CI, 95% confidence interval; FDR, false discovery rate.

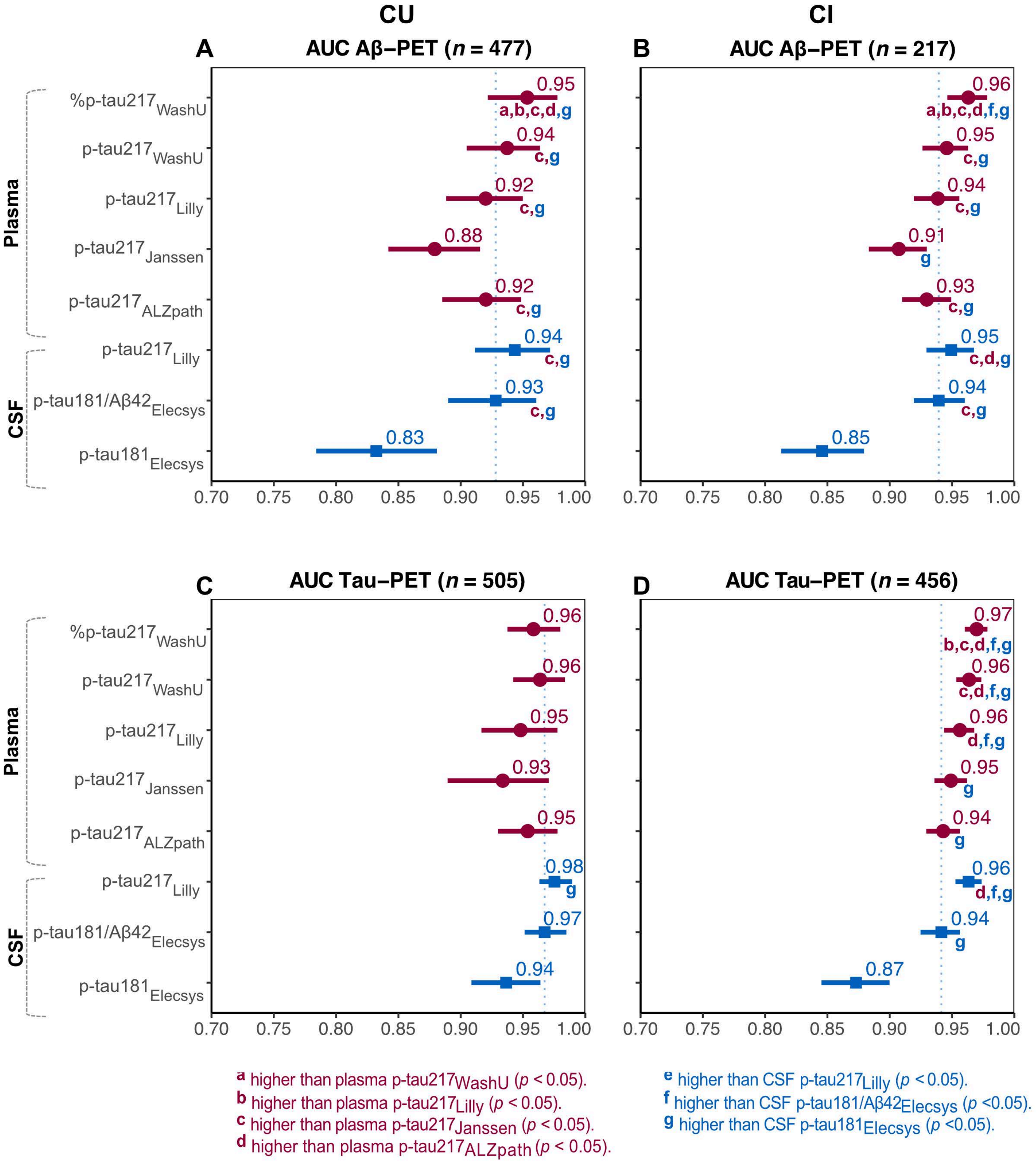

**Supplementary Figure 11. Accuracy comparisons in cognitively unimpaired and cognitively impaired.** The dashed line is drawn at CSF p-tau181/Aβ42$_{Elecsys}$ to facilitate comparing the other tests to the current approved FDA-approved test. Significant differences between assays were assessed through bootstrapping. In brief, a bootstrap distribution of the difference in accuracy of two different biomarkers was generated ($n$ = 1000), and their 95% CI. Then, p-values were derived from the distributed differences in values, which were subsequently FDR-corrected. *Abbreviations*: Aβ, amyloid-beta; CU, cognitively unimpaired; CI, cognitively impaired; 95% CI, 95% confidence interval; FDR, false discovery rate.

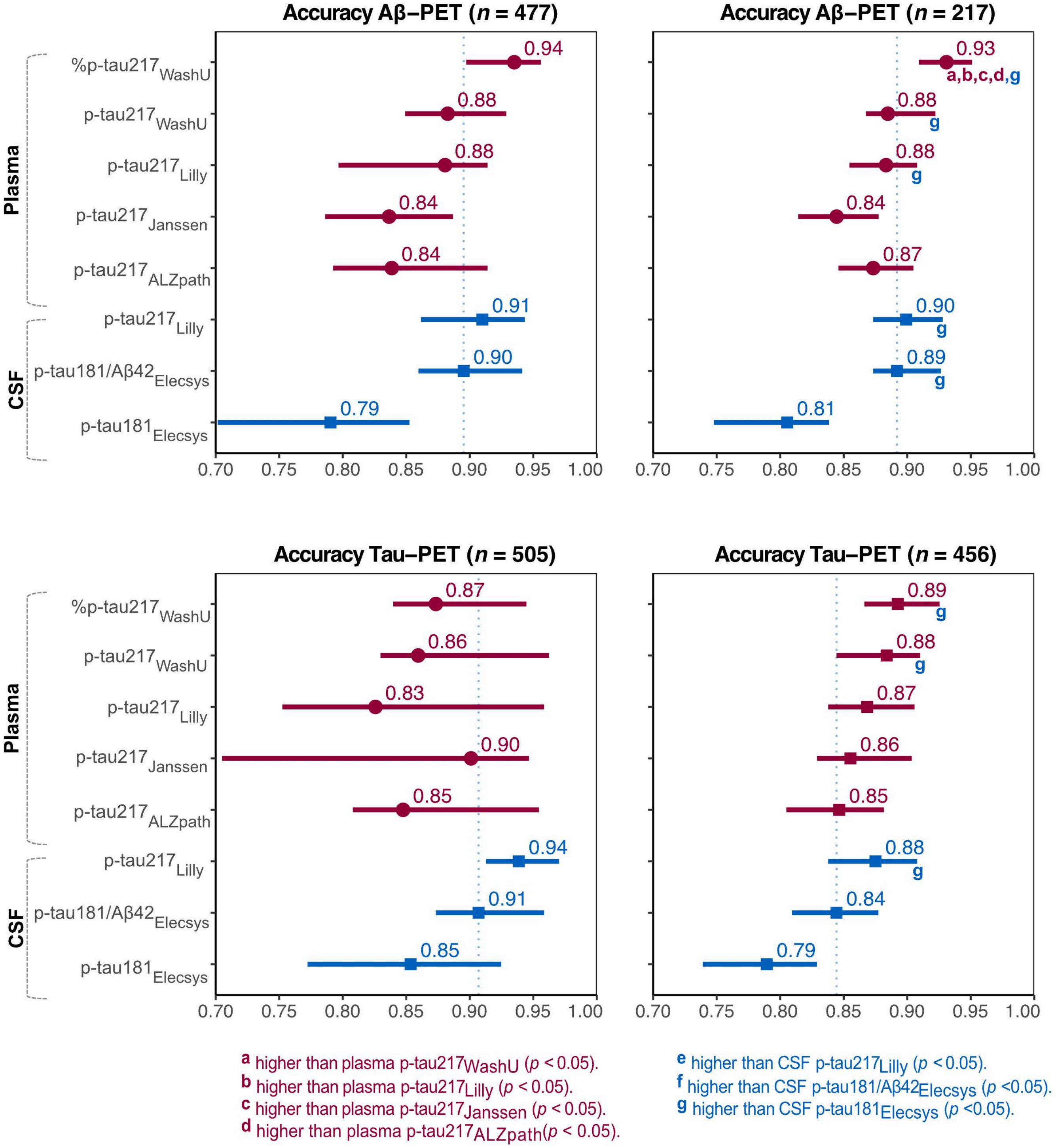

**Supplementary Figure 12. Cross-sectional PET $R^2$ comparisons in cognitively unimpaired and cognitively impaired.** The dashed line is drawn at CSF p-tau181/Aβ42$_{Elecsys}$ to facilitate comparing the other tests to the current approved FDA-approved test. Significant differences between assays were assessed through bootstrapping. In brief, a bootstrap distribution of the difference in $R^2$ of two different biomarkers was generated ($n$ = 1000 iterations). Then, p-values were derived from the distributed differences in $R^2$ values, which were subsequently FDR-corrected. Cross-sectional: PET SUVR ~ biomarker + age + sex + education. *Abbreviations*: Aβ, amyloid-beta; CI, confidence interval; FDR, false discovery rate.

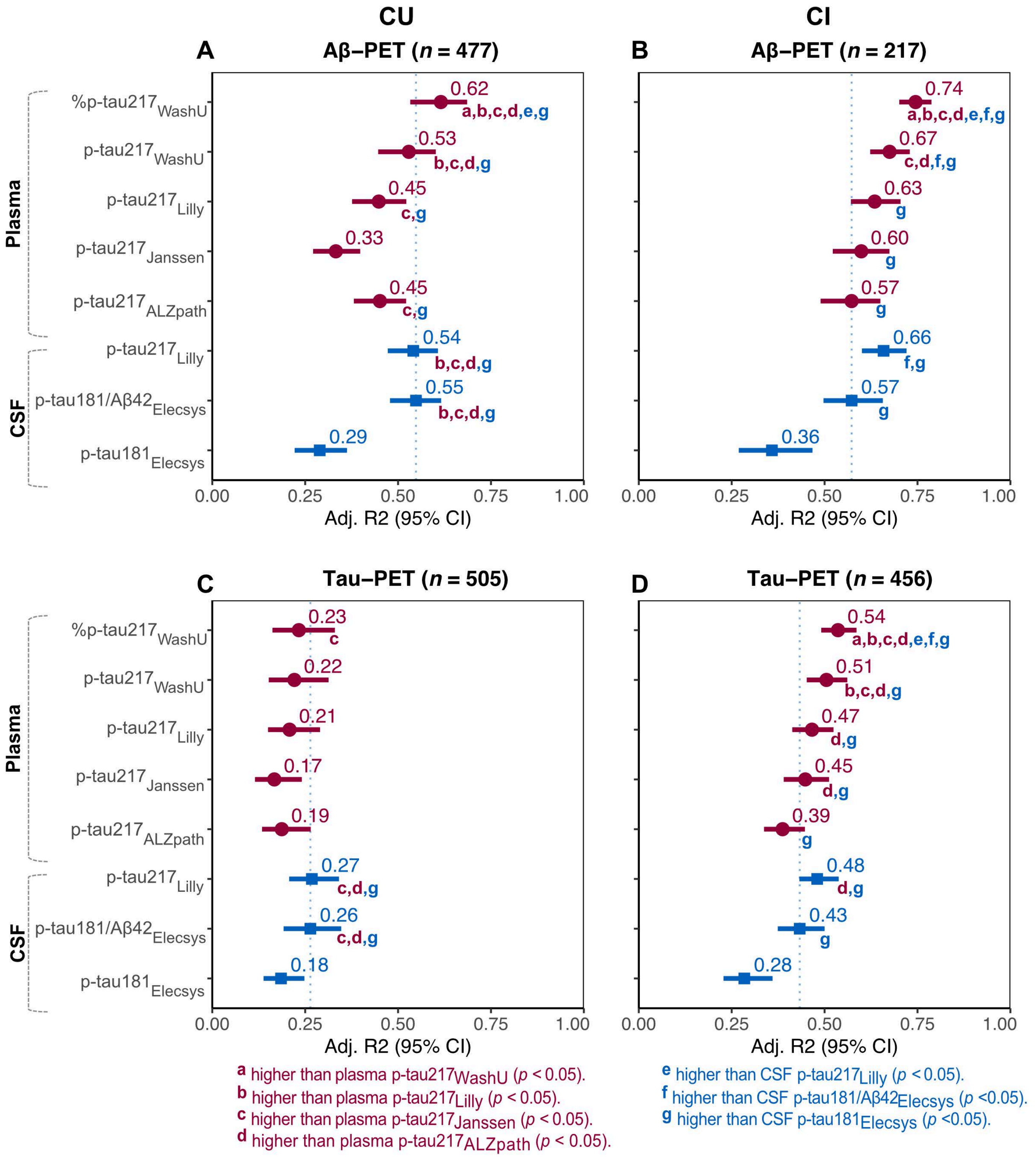

**Supplementary Figure 13. Longitudinal PET $R^2$ comparisons in cognitively unimpaired and cognitively impaired.** The dashed line is drawn at CSF p-tau181/Aβ42$_{Elecsys}$ to facilitate comparing the other tests to the current approved FDA-approved test. Significant differences between assays were assessed through bootstrapping. In brief, a bootstrap distribution of the difference in $R^2$ of two different biomarkers was generated (*n* = 1000 iterations). Then, p-values were derived from the distributed differences in $R^2$ values, which were subsequently FDR-corrected. The rate of change was obtained from linear mixed models, which was then used as the outcome in a linear regression model. Longitudinal: individual rate of change in PET SUVR ~ biomarker + age + sex + education. *Abbreviations*: Aβ, amyloid-beta; CI, confidence interval; FDR, false discovery rate.

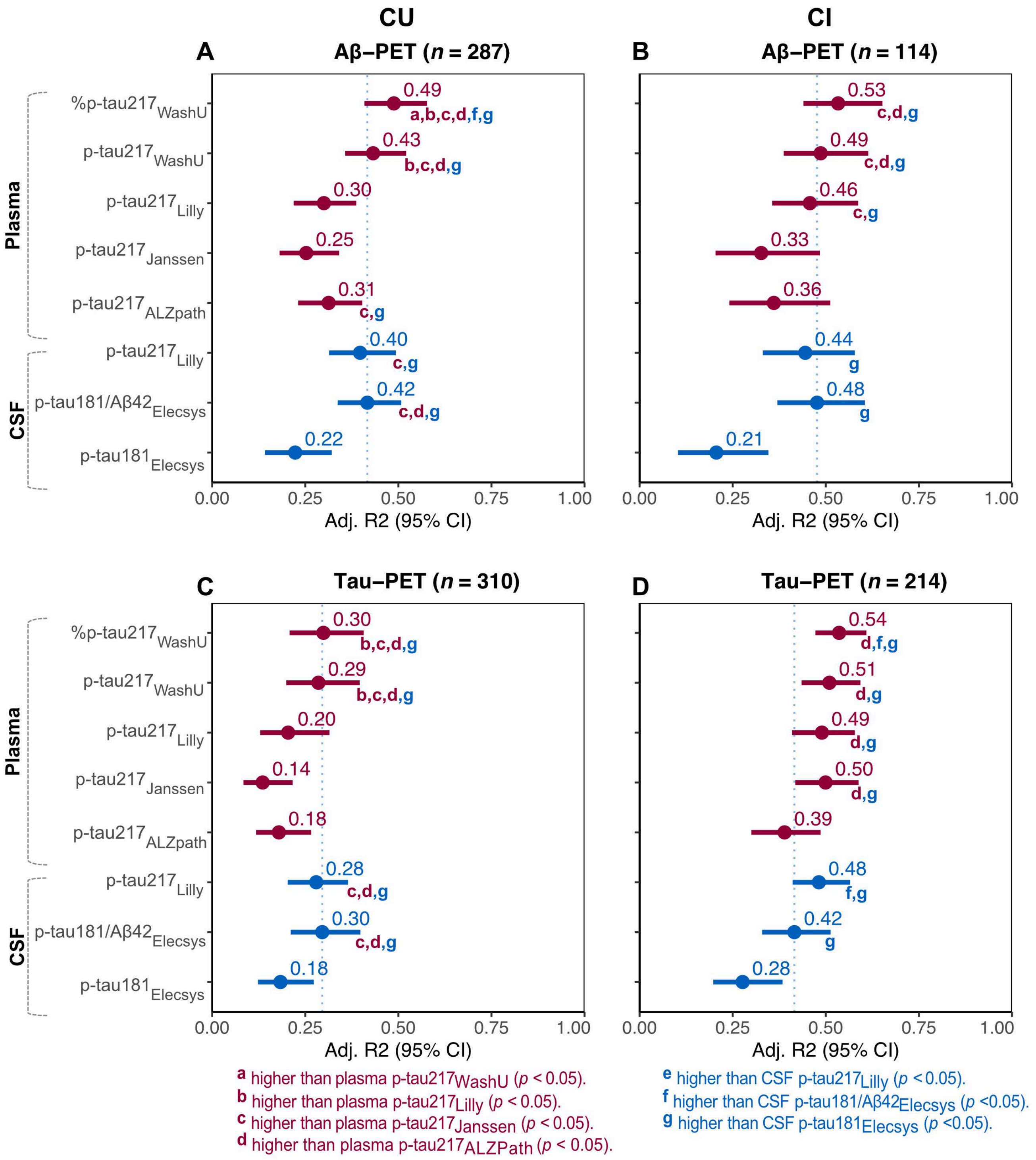

**Supplementary Figure 14. Cognition $R^2$ comparisons in cognitively unimpaired or cognitively impaired.** The dashed line is drawn at CSF p-tau181/Aβ42$_{Elecsys}$ to facilitate comparing the other tests to the current approved FDA-approved test. Significant differences between assays were assessed through bootstrapping. In brief, a bootstrap distribution of the difference in $R^2$ of two different biomarkers was generated ($n$ = 1000 iterations). Then, p-values were derived from the distributed differences in $R^2$ values, which were subsequently FDR-corrected. The rate of change was obtained from linear mixed models, which was then used as the outcome in a linear regression model. For the MMSE models, participants with non-AD dementia were excluded. MMSE models were only performed in CI individuals. For the mPACC models, participants with dementia were excluded. As the mPACC is intended for detection of early cognitive changes in Alzheimer's disease, mPACC models were only performed in CU individuals. Cross-sectional: cognition ~ biomarker + age + sex + education. Longitudinal: individual rate of change in cognition ~ biomarker + age + sex + education. *Abbreviations*: Aβ, amyloid-beta; CI, confidence interval; FDR, false discovery rate.

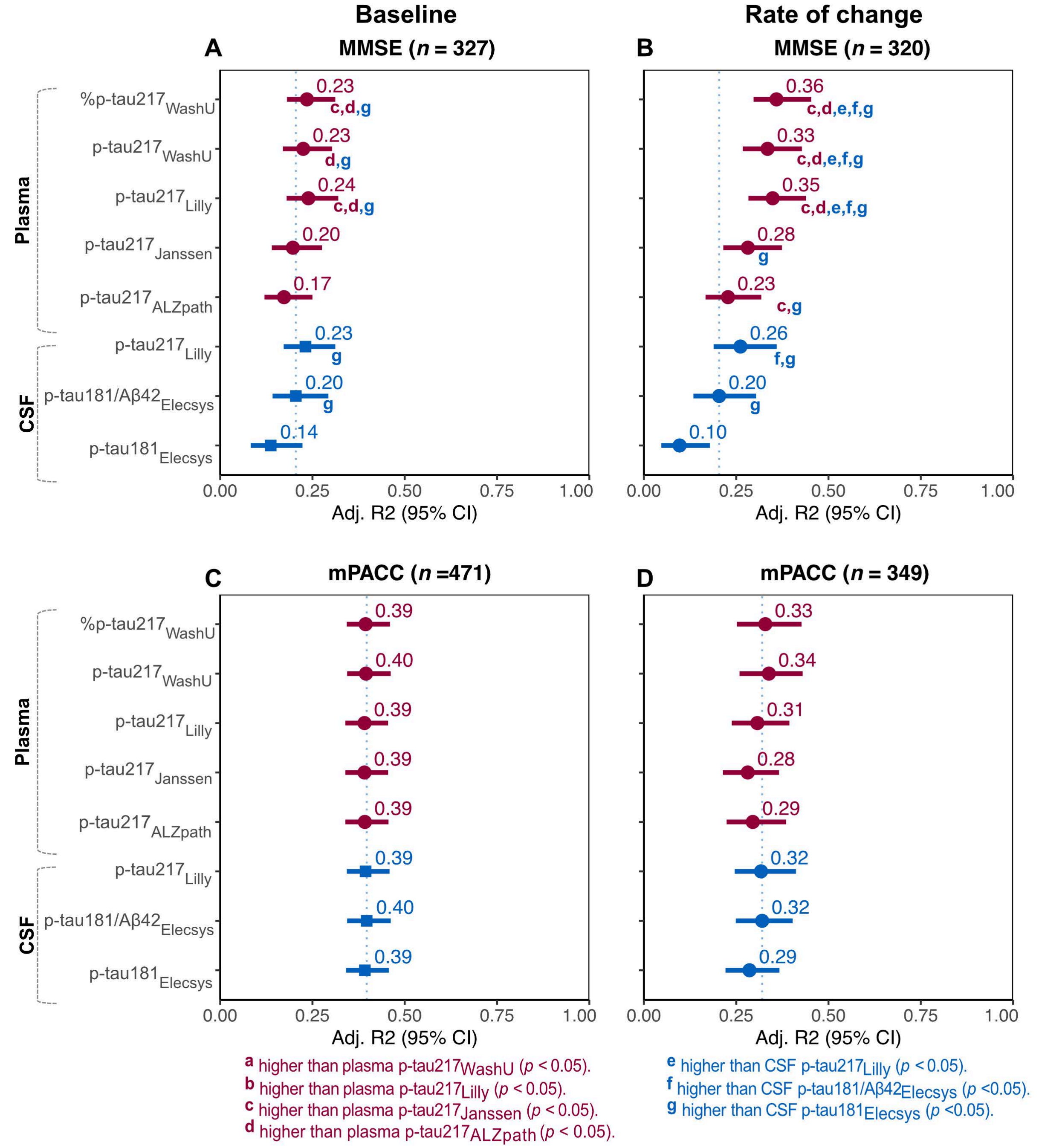

**Supplementary Figure 15. Replication including the NULISA assay.** Dots and squares represent the AUC, accuracy or R2, and bars represent 95%CI. The dashed line is drawn at CSF p-tau181/Aβ42$_{Elecsys}$, to facilitate comparing the other tests to the current approved FDA-approved test. The global cognitive composite was a composite of several cognitive tests, z-scored with CU Aβ- individuals as reference group. Significant differences between assays were assessed through bootstrapping. In brief, a bootstrap distribution of the differences in AUC, accuracy and $R^2$ of two different biomarkers was generated (*n* = 1000 iterations), and the 95% CI of the mean difference. Then, p-values were derived from the distributed differences in values, which were subsequently FDR-corrected. Linear model Aβ-PET: Aβ-PET ~ biomarker + age + sex. Linear model cognition: cognition ~ biomarker + age + sex + education. *Abbreviations*: Aβ, amyloid-beta; CI, confidence interval; CU, cognitively unimpaired; FDR, false discovery rate.

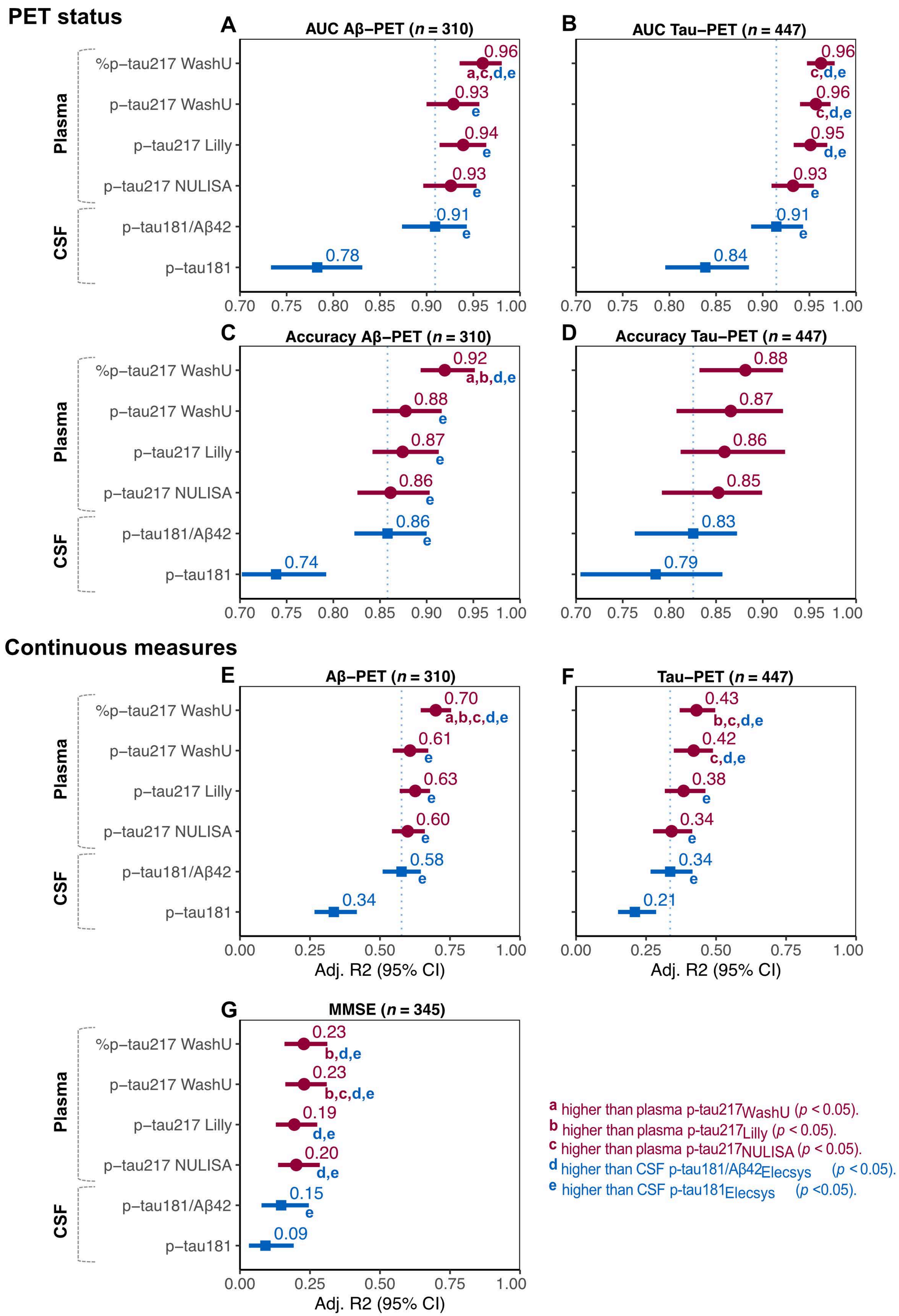
PET status
A AUC Aβ−PET (n = 310)
B AUC Tau−PET (n = 447)
C Accuracy Aβ−PET (n = 310)
D Accuracy Tau−PET (n = 447)
Continuous measures
E Aβ−PET (n = 310)
F Tau−PET (n = 447)
G MMSE (n = 345)
Plasma
CSF
%p−tau217 WashU
p−tau217 WashU
p−tau217 Lilly
p−tau217 NULISA
p−tau181/Aβ42
p−tau181
Adj. R2 (95% CI)
a higher than plasma p-tau217WashU (p < 0.05).
b higher than plasma p-tau217Lilly (p < 0.05).
c higher than plasma p-tau217NULISA (p < 0.05).
d higher than CSF p-tau181/Aβ42Elecsys (p < 0.05).
e higher than CSF p-tau181Elecsys (p <0.05).

**Supplementary Figure 16. Replication in the Knight ADRC cohort including the NULISA assay.** Dots and squares represent the AUC, accuracy or R2, and bars represent 95%CI. The dashed line is drawn at CSF p-tau181/Aβ42$_{Lumipulse}$, to facilitate comparing the other tests to the current approved FDA-approved test. The global cognitive composite was a composite of several cognitive tests, z-scored with CU Aβ- individuals as reference group. Significant differences between assays were assessed through bootstrapping. In brief, a bootstrap distribution of the differences in AUC, accuracy and $R^2$ of two different biomarkers was generated ($n$ = 1000 iterations), and the 95% CI of the mean difference. Then, p-values were derived from the distributed differences in values, which were subsequently FDR-corrected. Linear model Aβ-PET: Aβ-PET ~ biomarker + age + sex. Linear model global cognitive composite: cognition ~ biomarker + age + sex + education. *Abbreviations*: Aβ, amyloid-beta; CI, confidence interval; CU, cognitively unimpaired; FDR, false discovery rate.

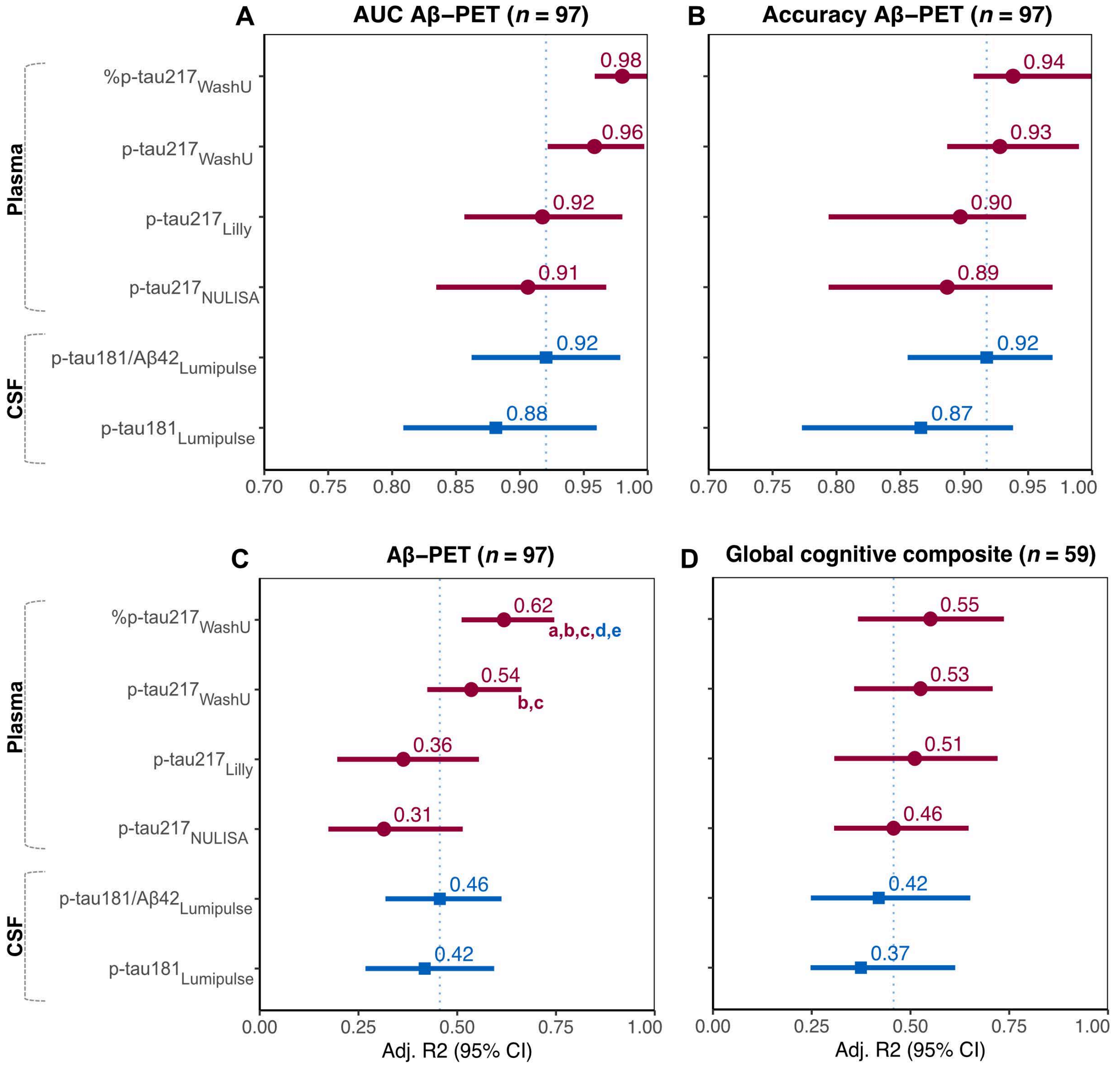


a higher than plasma p-tau217$_{WashU}$ ($p$ < 0.05).
b higher than plasma p-tau217$_{Lilly}$ ($p$ < 0.05).
c higher than plasma p-tau217$_{NULISA}$ ($p$ < 0.05).
d higher than CSF p-tau181/Aβ42$_{Lumipulse}$ ($p$ < 0.05).
e higher than CSF p-tau181$_{Lumipulse}$ ($p$ <0.05).
